# Prevalence and types of strabismus in cerebral palsy: A global and historical perspective based on a systematic review and meta-analysis

**DOI:** 10.1101/2024.01.23.24301684

**Authors:** Michael S. Herron, Lingchen Wang, Christopher S. von Bartheld

**Affiliations:** Center of Biomedical Research Excellence in Cell Biology, University of Nevada, Reno School of Medicine, Reno, Nevada, USA; School of Public Health, University of Nevada, Reno, Nevada, USA; Department of Physiology and Cell Biology, University of Nevada, Reno School of Medicine, Reno, Nevada, USA

**Keywords:** strabismus, cerebral palsy, esotropia, exotropia, global, ethnicity, geographic mapping

## Abstract

**Purpose:** Strabismus is more frequent in cerebral palsy (CP) than in the normal population, but reports differ how much it is increased. We here examined the global prevalence and types of strabismus in CP, whether esotropia or exotropia is more frequent, and whether the prevalence differs between ethnicities and/or country income levels, and between generations.

**Methods:** We compiled in a systematic review and meta-analysis the results of 147 CP studies that report the prevalence of strabismus or the ratio of esotropia to exotropia, and we conducted subgroup analyses for region (income level) and ethnicity. We performed a pooled analysis for the CP strabismus prevalence, and estimated the global number of CP cases with strabismus.

**Results:** The pooled prevalence of strabismus in CP is 49.8% in high-income countries and 39.8% in lower-income countries. We estimate the global number of strabismus cases in CP as 12.2 million, with 7.6 million males and 4.6 million females, based on current estimates of 29.6 million global CP cases. Esotropia is more frequent than exotropia in Caucasians, while exotropia is more frequent than esotropia in Hispanic and in some Asian and African populations. The strabismus prevalence in CP increases with increasing country income levels.

**Conclusion:** Generational changes in strabismus prevalence appear to reflect a transition of CP types and an increase in prevalence as countries attain higher income and more effective maternal health care. The distribution of esotropia and exotropia in CP patients largely reflects the horizontal strabismus type that is predominant in the subject’s ethnicity.

## Introduction

Strabismus is one of the most frequent co-morbidities of cerebral palsy (CP).^1–6^ While there is a consensus that strabismus is much more frequent in CP than in the normal, non-CP population (where it is estimated at 2-3%),^7,8^ reports of the prevalence of CP-associated strabismus differ considerably, ranging from 15% or less^9–13^ to over 90%.^14–16^ The prevalence of strabismus in CP differs not only between studies, but may differ between geographic regions or ethnicities. The only previous systematic review of strabismus in CP^6^ estimated the prevalence of strabismus at 48%, but Caucasians in high-income countries were over-represented, while Africans and Asians in lower-income countries were under-represented.

It is not known how many cases of CP with strabismus exist worldwide. Some of this uncertainty is due to controversies about the global prevalence of CP. CP is well-studied in Caucasian populations, in high-income countries, where the prevalence is about 2 per 1,000,^17–20^ but the CP prevalence in lower-income countries (which is the large majority of the world population) is uncertain, with estimates ranging between 2 and 10 per 1,000.^5,16,18,20–32^ Since CP types differ between developing and high-income countries,^26,33–35^ and most authors conclude that the type of CP associates with strabismus prevalence,^5,6,10,11,28,33,35–47^ it needs to be explored whether socioeconomic factors and/or ethnicity are significant variables for the strabismus prevalence among CP patients.

Another issue of contention is whether esotropia (ET) or exotropia (XT) is more frequent in CP. Initial studies, most of them examining populations of European ancestry, report more ET than XT, although some studies noted a lower ET/XT ratio in CP than in the general population.^15,48–53^ Several of the more recent studies, many of them from East Asia and the South of the Indian subcontinent, reported more XT than ET in CP patients.^54–59^ It has not been established whether the type of horizontal strabismus in CP is associated with the type of the underlying brain lesion, or whether it rather reflects the type of horizontal strabismus generally seen within the same ethnicity. We sought to answer the following questions:

1. What is the global prevalence of strabismus in people with CP? How much more frequent is it compared to the non-CP population? Can the global number of CP cases with strabismus be estimated?
2. Is the prevalence of strabismus in CP the same in Caucasians as in other ethnicities or regions (lower-vs high-income countries)?
3. Is the ET/XT ratio in CP significantly different between ethnicities?
4. Does the type of brain lesion in CP influence the type of horizontal strabismus (ET or XT), or does the direction of deviation reflect the type of horizontal strabismus that is predominant in the same ethnic population?
5. Are there longitudinal trends of strabismus prevalence in CP over generations?

To resolve these issues, we performed a systematic review and meta-analysis of strabismus in CP that was initially posted as a preprint.^60^ Our review provides a comprehensive compilation of relevant studies, and we map their geographic distribution. We offer a global and historical perspective by exploring the role of ethnicity and socioeconomic factors.

## Materials and Methods

### Search Strategy

For our systematic review of the literature, we adhered to the PRISMA guidelines.^61^ Reports of studies were identified through a search of three databases: Google Scholar, PubMed, and CNKI (China National Knowledge Infrastructure), with unrestricted years. We used the keywords “cerebral palsy”, “strabismus”, “esotropia” and “exotropia” in Google Scholar, and “cerebral palsy” and “strabismus”, as well as “cerebral palsy” and “squint” in PubMed. Only English terms were used for the search strategy of the first two databases. We searched the CNKI database using the Chinese words for “cerebral palsy” and “strabismus” as key words. Reference lists from eligible articles were examined to find additional relevant studies. Studies were also identified by searching papers that cited relevant sources. All titles were screened, and when potentially relevant, the abstract was evaluated to decide whether the paper should be obtained for full-text reading to verify eligibility (Fig. 1). We failed to obtain an abstract and full text in two cases (2/708).

**Fig. 1.**
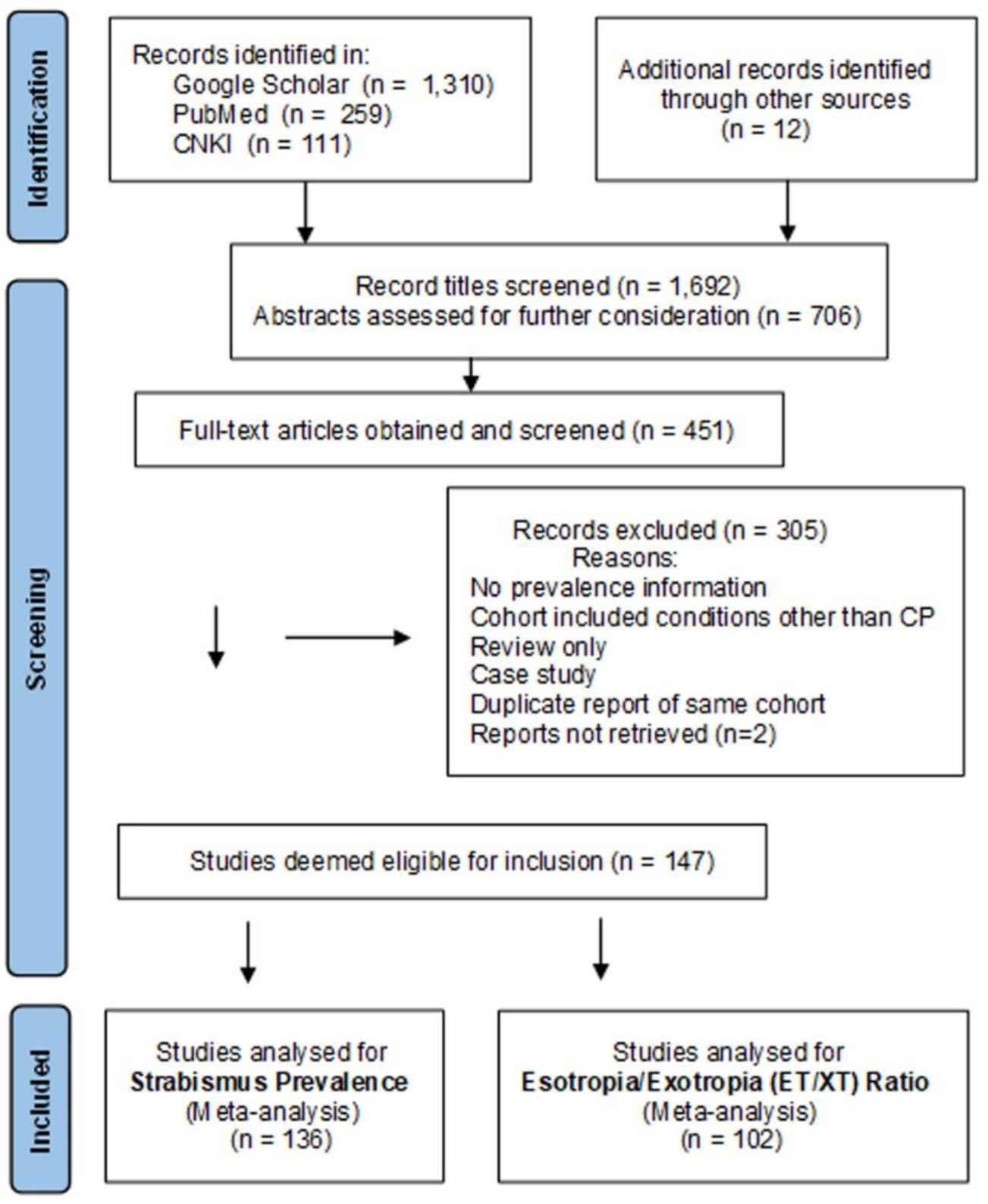
Flowchart of the Literature Search and Screening Strategy.

### Inclusion/Exclusion Criteria

To be eligible for the systematic review, studies had to report the numerical prevalence of strabismus in humans with cerebral palsy (CP) and/or provide the ratio of esotropia vs exotropia in a CP cohort. CP was defined as a “disorder of movement and posture due to a defect or lesion of the immature brain”, a definition that was subsequently refined and expanded.^62,63^ We excluded studies that reported on children with various other causes of visual impairment, unless they reported specific data for a CP cohort. We excluded disability conditions that can be similar to CP, such as premature birth, hydrocephalus, microcephalus, meningitis, developmental delay, Down syndrome, epilepsy, or autism spectrum disorders. We also excluded reviews only, case studies, abstracts at meetings when later published as a peer-reviewed paper, and papers focused on surgical outcomes. We included 54 studies on Caucasians (most of them in Europe, North America and Australia),^9,10,14,36–42,44,48–53,64–101^ 11 studies from the Middle East,^5,16,23,102–109^ 9 on Hispanics (most of them from Latin America),^15,110–117^ 41 from East Asia,^12,13,43,46,56,58,118–152^ 23 from South Asia,^33,35,45,47,54,57,153–169^ and 5 from Africa^11,170–174^ (Supplemental Table 1).

### Data Extraction and Analyses

Data were extracted by using pre-designed tables, including date of publication, first author name, country, geographic region, cohort size, number of cases of strabismus, and, when available, age range and the type of strabismus: horizontal vs. vertical, and among the horizontal strabismus, how many cases of esotropia, and how many cases of exotropia. We compiled information about gender distribution in the cohort and in strabismus cases when such data was reported. The percentage of strabismus cases was calculated from the number of cases per cohort. We conducted subgroup analyses between ethnicities, as well as between high-income and lower-income countries. Income levels were defined by the GDP as per the World Bank Report (1987 to 2022).^175^ Because of ethnic differences between populations, the prevalence for each major ethnicity (European ancestry, African, Middle East, East Asian, South Asian, Latino/Hispanic) was estimated separately and weighted by population size to generate an overall estimate of the current global strabismus prevalence in CP. This was also necessary to prevent bias: the largest fraction of available studies examined people with European ancestry (in high-income countries). There were sufficient data for Caucasians, South Asians, and East Asians to assess generational (longitudinal) trends.

### Statistical Analyses

The primary purpose of our meta-analysis was to generate a more precise and reliable estimate of the prevalence of strabismus among people with CP and to predict the global number of CP cases with strabismus. Pooled analyses were performed for strabismus prevalence in CP and the ET/XT ratio. The heterogeneity among studies was evaluated by Cochran’s Q test and the I^2^ index.^176,177^ The random-effect models were used to conservatively diminish the heterogeneity between studies.^177^ The study weights were obtained based on the DerSimonian-Laird method.^177^ A continuity correction of 0.5 was applied to studies with zero cases.^176^ Subgroup pooled analyses were conducted by region/ethnicity, separately for prevalence and for the ET/XT ratio, to assess differences between Caucasians and other ethnicities. Meta-regression analyses were performed to test associations between risk factors and study outcomes. The risk of publication bias was evaluated using funnel plots and Egger’s test.^178^ The significance level was set to 0.05. All meta-analyses were performed using the Stata SE 16.0 software (StataCorp, TX, USA).

## Results

Our analyses are based on 147 studies, with 136 of them reporting the prevalence of strabismus in CP (total cohort number: 21,449), 102 partially overlapping studies reporting the esotropia/exotropia (ET/XT) ratio in CP cases (total cohort number: 13,613), and 90 partially overlapping studies reporting the gender distribution in the CP cohorts (total cohort number: 15,728). Altogether, these studies examined 21,753 CP cases in cohorts from 29 countries on five continents.

### Global prevalence of CP and estimate of the total number of CP cases worldwide

While the prevalence of CP in developed (high-income) countries is well-established at about 2/1,000,^2,19,20,24,26,44,49,64,71,179^ there is less information about the prevalence of CP in lower-income countries.^17,25,27–31^ Initially, it was assumed that the CP prevalence was similar between developed and developing countries.^17–19,24^ Recent studies have revealed a much higher prevalence of CP in many lower-income countries.^20,25,26,30,31^ For example, the prevalence of CP per 1,000 in India was reported as 2.8,^180^ in rural Uganda it is 3,^27^ in Bangladesh 3.4,^28^ in Egypt 3.6,^181^ in Mexico 4.4,^32^ in Turkey between 4.4 and 5.6,^22,182^ and in rural South Africa as 10.^21,25^ These prevalences are thought to be underestimates.^20,28^ Some authors noted that the prevalence of CP in lower-income countries is twofold larger than in high-income countries.^20,26,28^ Very recent studies, based on new data and statistical modeling, estimated the prevalence and total number of global CP cases significantly greater, at about 6 per 1,000,^20,27,28,30^ equivalent to about 50 million global cases.^29,31^ However, the statistical modeling appears to include cases with motor dysfunction similar to CP, but not strictly with a CP diagnosis.^29^ The much higher CP prevalence in lower-income countries has important implications for the estimates of the global prevalence of CP.

In our approach to estimate global CP numbers, we applied a prevalence of 2/1,000 for high-income countries, and 4/1,000 for lower-income countries (distinction according to The World Bank).^175^ We estimate the CP cases in high-income countries (population of about 1.2 billion) to be 2.4 million (1.2 billion x 0.2%), and in lower-income countries (population of about 6.8 billion) to be 27.2 million (6.8 billion x 0.4%), which combines for a global total of 29.6 million CP cases. This estimate is intermediate between the previous, too low notion of 18 million global CP cases,^183^ and the likely too high 50 million global cases.^29^

### Global prevalence of strabismus in CP: estimate of total numbers of CP cases with strabismus

The geographic distribution of studies reporting the prevalence of strabismus in CP is shown in Fig. 2A, with the prevalence indicated by a color gradient and the size of the cohort reflected by the size of the circles. The prevalence of strabismus in CP seems lower in some regions (Africa, for example) when compared with other regions. We therefore performed subgroup analyses to determine whether socioeconomic conditions or ethnicity are significant factors. Subgroup analyses for the major ethnicities were not significantly different, except for Caucasians vs. East Asians (p=0.001, Fig. 3A). Subgroup analysis by country income level showed a significantly higher strabismus prevalence for high income (49.8%, 95% confidence interval CI=44.3–55.2%) than for lower income (39.8%, CI=34.4–45.1%, p=0.004). The global strabismus prevalence was 41.3% when adjusted for the population size of high-income countries (1.2 billion) and lower-income countries (6.8 billion, Table 1A). When the differences in CP cases between regions as well as the differences between prevalence of strabismus in CP cases are taken into account, we can use the above estimate of the global number of CP cases (29.6 million) to calculate the total number of CP cases with strabismus worldwide – as 12.2 million (29.6 x 41.3%).

**Fig. 2.**
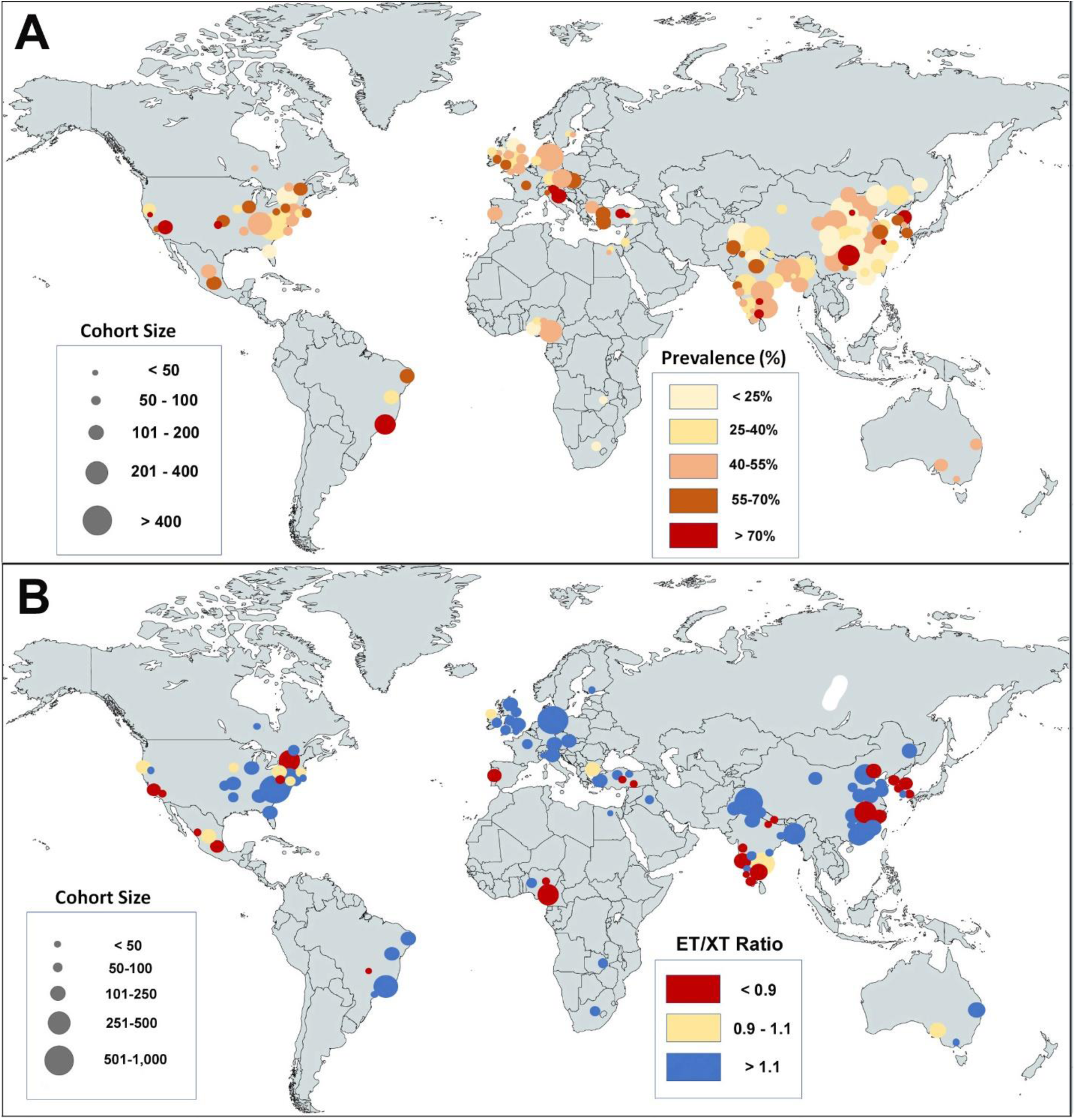
**A, B.** World maps showing the distribution of studies reporting the prevalence of strabismus in cerebral palsy (**A**) and the esotropia/exotropia (ET/XT) ratio in cerebral palsy (**B**). The cohort size is indicated by the circle size and the prevalence or ratio is indicated by the color gradient. In **A**, note that lower-income countries (Africa and India) have a somewhat lower prevalence than high-income countries. In **B**, the ET/XT ratio below 0.9 is indicated in red, a nearly equal distribution (0.9 to 1.1) is indicated in tan, and a ratio above 1.1 is indicated in blue. Note that regions with Caucasian populations mostly have a high ET/XT ratio, while some populations in Africa, East Asia, Southern India and some Hispanic populations have a low ET/XT ratio.

**Fig. 3.**
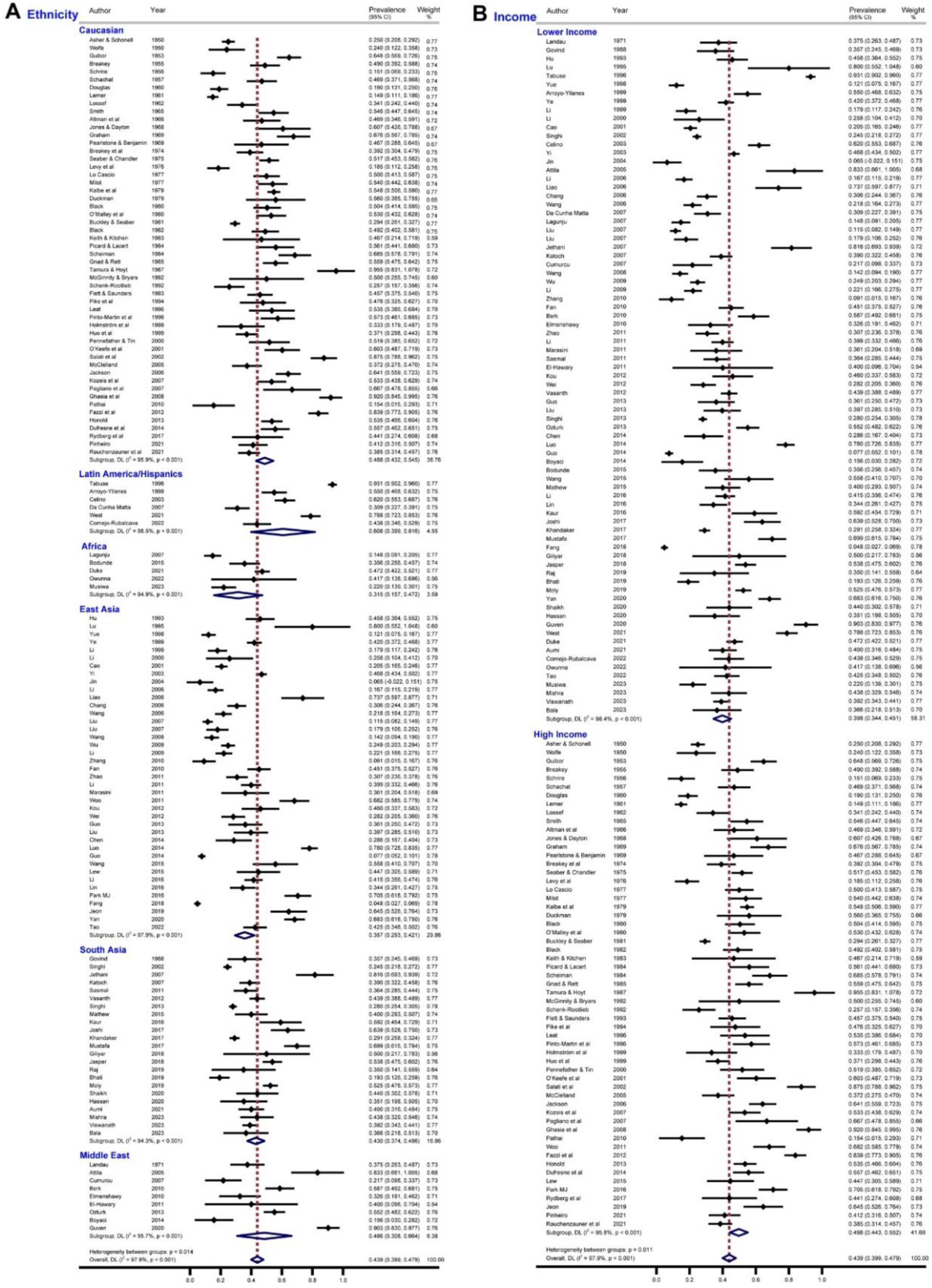
**A, B:** Forest Plots of the Prevalence of Strabismus in Cerebral Palsy. **A.** Sorted by ethnicity. **B.** Sorted by lower-income countries vs high-income countries – a statistically significant difference (p=0.004, see Supplemental Fig. 2A). Note the shift in high-income countries towards a higher prevalence of strabismus. The diamonds show the pooled prevalence for the listed studies.

**Table 1.**
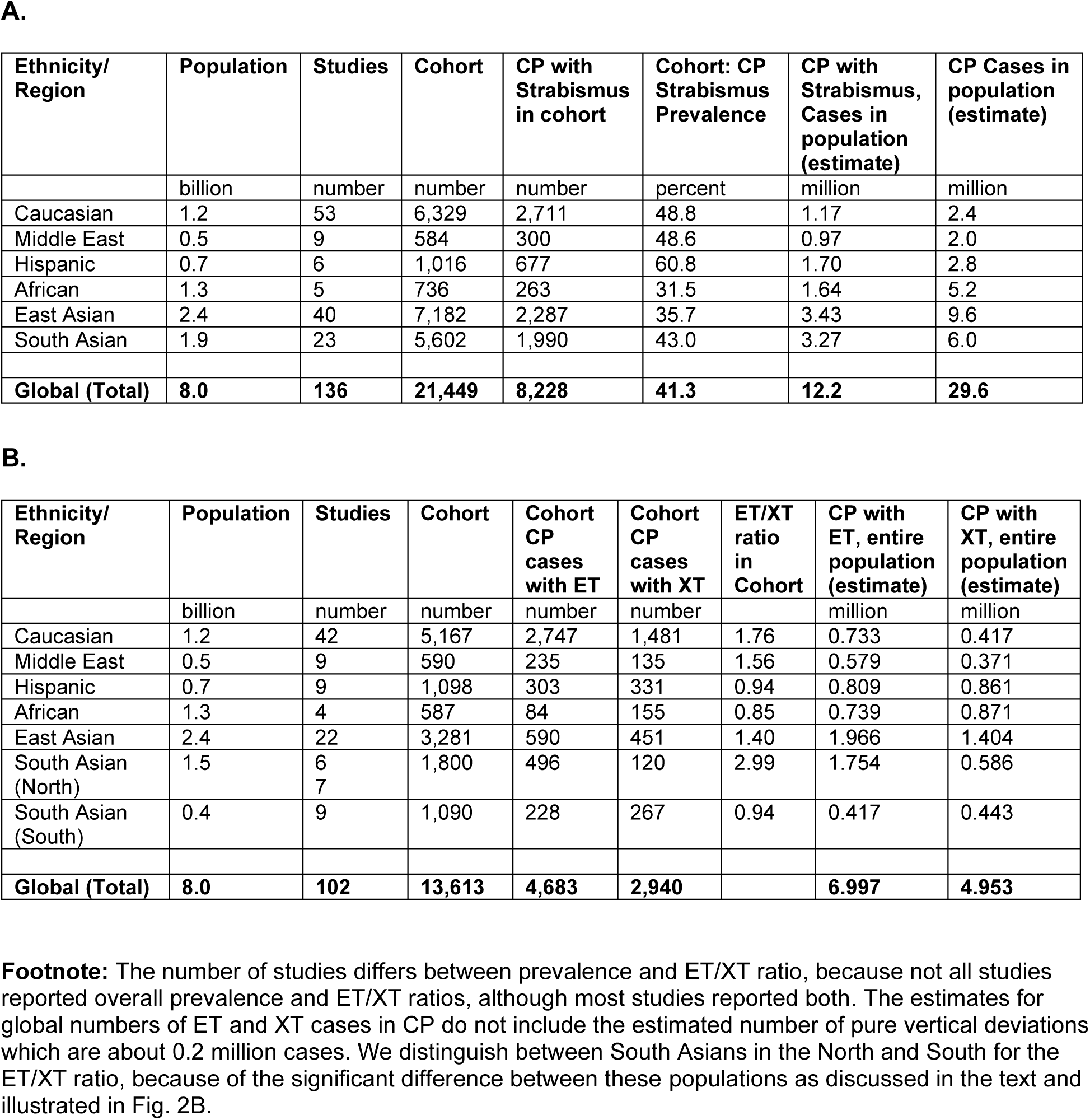
Prevalence of strabismus in cerebral palsy (CP): Data and estimates of numbers (**A**). Esotropia (ET) and exotropia (XT) in CP: Data and estimates of numbers (**B**).

| Ethnicity/<br>Region | Population | Studies | Cohort | CP with<br>Strabismus<br>in cohort | Cohort: CP<br>Strabismus<br>Prevalence | CP with<br>Strabismus,<br>Cases in<br>population<br>(estimate) | CP Cases in<br>population<br>(estimate) |
| --- | --- | --- | --- | --- | --- | --- | --- |
|  | billion | number | number | number | percent | million | million |
| Caucasian | 1.2 | 53 | 6,329 | 2,711 | 48.8 | 1.17 | 2.4 |
| Middle East | 0.5 | 9 | 584 | 300 | 48.6 | 0.97 | 2.0 |
| Hispanic | 0.7 | 6 | 1,016 | 677 | 60.8 | 1.70 | 2.8 |
| African | 1.3 | 5 | 736 | 263 | 31.5 | 1.64 | 5.2 |
| East Asian | 2.4 | 40 | 7,182 | 2,287 | 35.7 | 3.43 | 9.6 |
| South Asian | 1.9 | 23 | 5,602 | 1,990 | 43.0 | 3.27 | 6.0 |
| **Global (Total)** | **8.0** | **136** | **21,449** | **8,228** | **41.3** | **12.2** | **29.6** |

**B.**
| Ethnicity/<br>Region | Population | Studies | Cohort | Cohort<br>CP<br>cases<br>with ET | Cohort<br>CP<br>cases<br>with XT | ET/XT<br>ratio<br>in<br>Cohort | CP with<br>ET, entire<br>population<br>(estimate) | CP with<br>XT, entire<br>population<br>(estimate) |
| --- | --- | --- | --- | --- | --- | --- | --- | --- |
|  | billion | number | number | number | number |  | million | million |
| Caucasian | 1.2 | 42 | 5,167 | 2,747 | 1,481 | 1.76 | 0.733 | 0.417 |
| Middle East | 0.5 | 9 | 590 | 235 | 135 | 1.56 | 0.579 | 0.371 |
| Hispanic | 0.7 | 9 | 1,098 | 303 | 331 | 0.94 | 0.809 | 0.861 |
| African | 1.3 | 4 | 587 | 84 | 155 | 0.85 | 0.739 | 0.871 |
| East Asian | 2.4 | 22 | 3,281 | 590 | 451 | 1.40 | 1.966 | 1.404 |
| South Asian<br>(North) | 1.5 | 6<br>7 | 1,800 | 496 | 120 | 2.99 | 1.754 | 0.586 |
| South Asian<br>(South) | 0.4 | 9 | 1,090 | 228 | 267 | 0.94 | 0.417 | 0.443 |
| **Global (Total)** | **8.0** | **102** | **13,613** | **4,683** | **2,940** |  | **6.997** | **4.953** |

### Frequencies, numbers and ratios of esotropia and exotropia cases in CP

The large majority of strabismus cases in CP are horizontal deviations, either esotropia (ET) or exotropia (XT), with a much smaller percentage of strabismus cases that have an exclusive vertical deviation (1.8%, CI=1.2–2.4%), based on 5,092 CP cases with information on hypertropia, Supplemental Table 1). The predominance of horizontal over vertical deviations is similar in non-CP and in CP populations. In the rest of our review, we will focus on horizontal strabismus. Most studies (74.5%) report more ET than XT in cases of CP (Fig. 2B; Supplemental Table 1). Populations with more ET than XT include Caucasians in Europe and North America, and most of the populations in the Middle East, South Asia, and East Africa. Populations with more XT than ET include parts of East Asia, the South of the Indian subcontinent, West Africa, and Hispanics in Central America (Figs. 2B; 4). Differences in the ET/XT ratio reached significance in Caucasians vs Hispanics (p=0.036) and was borderline for South Asians (South) vs Caucasians (p=0.062). Based on the estimate of the global prevalence of CP (see section above), we calculated the total number of CP cases with ET or with XT. Weighted by population size, the total global number of CP cases with ET is 7.0 million, and the total global number of CP cases with XT is 5.0 million (Table 1B).

### Gender distribution of strabismus in CP

Among our eligible CP studies, 89 report the gender distribution in the cohort, but only two of these studies also report the gender of the cases with strabismus.^166,167^ In CP cohorts, males nearly always exceed females, overall by a factor of about 1.5 to 1.^184^ The same is true for “our” CP cohorts – the ones that have been examined for strabismus (n=15,728), with 9,808 males and 5,920 females), which is a male/female ratio of 1.66 to 1 (7.6 million males and 4.6 million females). Two of these studies disclosed gender among strabismus cases (total cohort of 484)^166,167^; they indicate that there is no gender difference – males and females appear to contribute equally to the strabismus cases in the CP cohort, with the male and female prevalence of strabismus not being significantly different (p=0.440). In other words, there are more males with CP-associated strabismus than females, but only because more males than females have CP.

### Longitudinal analysis: strabismus prevalence in CP has increased over decades

To determine whether the prevalence of CP-associated strabismus has changed between generations, we examined the prevalence data in three ethnicities: Caucasians, East Asians, and South Asians (Supplemental Table 1). There was a significant trend in Caucasians from about 35% to 55% prevalence of strabismus in CP between 1950 and 1995, and the trendline slightly decreased from about 55% to 50% between 1995 and 2022 (Fig. 5A). The data for East Asians also showed an increasing trendline (Fig. 5B), from about 28% to 37%, but a nearly flat trendline for South Asians (Fig. 5C). These trends are consistent with the notion that strabismus prevalence in CP is associated with socioeconomic factors (see Discussion).

## Discussion

An association between CP and strabismus was first noted in the middle of the 19^th^ century; for a review of the early literature, see Smith.^1^ In the second half of the 20^th^ century, a series of more detailed studies from Europe and North America reported on the prevalence of strabismus in CP (Supplemental Table 1). Until the year 2000, most studies (39/49=79.6%) were carried out on Caucasian populations, thereby creating a Eurocentric bias. More recent studies examined populations from other regions of the world, notably in South Asia, East Asia and the Middle East (Table 1A; Supplemental Table 1), allowing to gain a better global perspective.

### Global prevalence of strabismus in CP

We estimate the total number of CP cases worldwide at 29.6 million (Table 1A). Given the incomplete ascertainment of CP cases as well as survival bias in developing countries, our estimate of 29.6 million CP cases globally may be an underestimate.^20,25,27,28,34^ How many of these CP patients have strabismus? The only previous systematic review estimated a 48% strabismus prevalence in CP which was based on 17 studies, with a combined cohort of 1,734.^6^ We find a lower global prevalence, of 41.3%. Our estimate is based on a much larger number of CP cases (21,449 cases), nearly 13-fold larger than the cohort size in the previous estimate.

The higher prevalence estimates^3–6,185^ are likely due to bias towards Caucasian populations in high-income countries.

The CP prevalence is known to increase with lower socioeconomic status.^18,25,28,34,186–188^ The recent studies from Africa and from South and East Asia are consistent with this notion. We show that lower-income countries (LICs) have a lower prevalence of CP-associated strabismus than high-income countries (HICs) (Fig. 3B). The key to understanding this surprising finding likely is that the timing of the brain insult and the mechanism of CP (and possibly strabismus) differ between LICs and HICs (Supplemental Fig. 2B). In LICs, the majority of CP cases are caused by peri- and postnatal events: asphyxia during delivery, or post-partum infection, while in HICs, the majority of CP cases are due to preterm births – when the immature brain is more vulnerable to lesions of the visual pathways.^26,28,33,34,185,189,190^ Additional factors complicate this basic pattern, in that very preterm babies rarely survive in LICs, but are more likely to survive in HICs, and that severe cases of CP are also more likely to survive in HICs, while in LICs, many may die prior to the age of CP diagnosis.^24,27,28,34^ This explanation is consistent with the observed data of a lower CP prevalence in HICs, but a higher prevalence of strabismus in CP cases in HICs than in LICs.

The trendlines of strabismus prevalence in CP (Fig. 5) are consistent with the prediction that developing countries will undergo similar shifts in CP types as developed countries did several decades ago – when more effective maternal health care systems were implemented. With an increasing number of studies from Asian, African and Hispanic populations, it now is possible to estimate the global prevalence of CP-associated strabismus and the global number of such cases. Accordingly, we estimate about 12.2 million such cases worldwide. This allows us to calculate how many cases of strabismus, due to CP, need to be added to the number of strabismus cases in the normal (non-CP) population. The normal (non-CP) population is thought to have a strabismus prevalence of 2-3%.^7,8^ Since most of the epidemiological studies rely on surveys of normal schools,^8,191^ many of the CP-associated strabismus cases (the ones with more severe CP) would have been missed. The ∼12.2 million CP strabismus cases comprise 6% of the total global number of strabismus cases (2-3% of a population of 8 billion people translates to 160-240 million cases).^7,8^

### Variation between studies

There is considerable variability in the prevalence of CP-associated strabismus between studies, ranging from less than 15% to over 90%, I^2^=97.9 (Table 1A; Fig. 3A,B; Supplemental Table 1). Multiple factors, not mutually exclusive, may contribute to this variability. First, the severity of CP differs between cohorts, and severity of CP is associated with strabismus prevalence. When cohorts primarily comprise mild cases of CP, they tend to have a lower prevalence of strabismus, while cohorts with severe cases have a higher prevalence of strabismus^9,37,192^. Second, the types of brain lesions differ between studies and between populations, due to differences in maternal health care as mentioned above. Third, ethnicity may be a significant variable, due to differences between populations in CP prevalence.^193,194^ Whether socioeconomic factors can fully account for such ethnic differences is controversial.^193,195^ Ethnicity may affect the type of strabismus (ET vs XT, see below), as well as the overall prevalence of strabismus.^142^ Fourth, methodological issues likely play a role, especially the criteria used to identify and diagnose CP cases which varies between countries and studies.^19,29,34,188^ Finally, there likely is natural variation between populations and also between risk factors for both CP and strabismus that may change over time – e.g., maternal smoking is a major risk factor for strabismus,^196,197^ regardless whether the offspring has CP or not. All of these factors likely contribute to the observed variation between studies.

### Is strabismus prevalence associated with CP types?

Strabismus prevalence is associated with the severity of CP, visual impairment, and also with the extent of intellectual capacity.^3,5,37,38,45,46,48,58,70,115,153,159,198–202^ Relatively few authors disagree with this conclusion.^39,74,101^ Some authors noted that strabismus is most frequent in spastic tetraplegia (or triplegia) and somewhat less in diplegia, still less in hemiplegia, and rare in athetoid or ataxic CP,^5,11,36,39,40,45,47,64,70,101,105,149,163^ while others disagree with this notion.^15,41–43,52,56,67,78,100,111,203^ Authors concur that there is no association between the *type* of strabismus (esotropia, exotropia, hypertropia) and the *type* of CP.^15,46,51,52,67,100,114^

### The esotropia/exotropia (ET/XT) ratio in CP

Our work has revealed differences between populations in how prevalent ET and XT are in CP (Figs. 2B, 4). Why do Caucasians, North Indians and people in the Middle East have more ET than XT, while populations in West Africa, Hispanics in Central America, and South Indians have more XT than ET? CP increases the likelihood of strabismus, but whether the strabismus will be ET or XT depends largely on the ethnicity, presumably due to differences in orbital anatomy, as first proposed by Holm^204^ and Waardenburg.^205^ The ET/XT ratio appears to be changed in CP (closer to 1), as has been noted by several authors for Caucasians^15,48,50–53^ and for East Asians.^56^ It seems that CP “tempers” the extremes and makes the ET/XT ratio more balanced than it is in the non-CP population (of the same ethnicity).

**Fig. 4:**
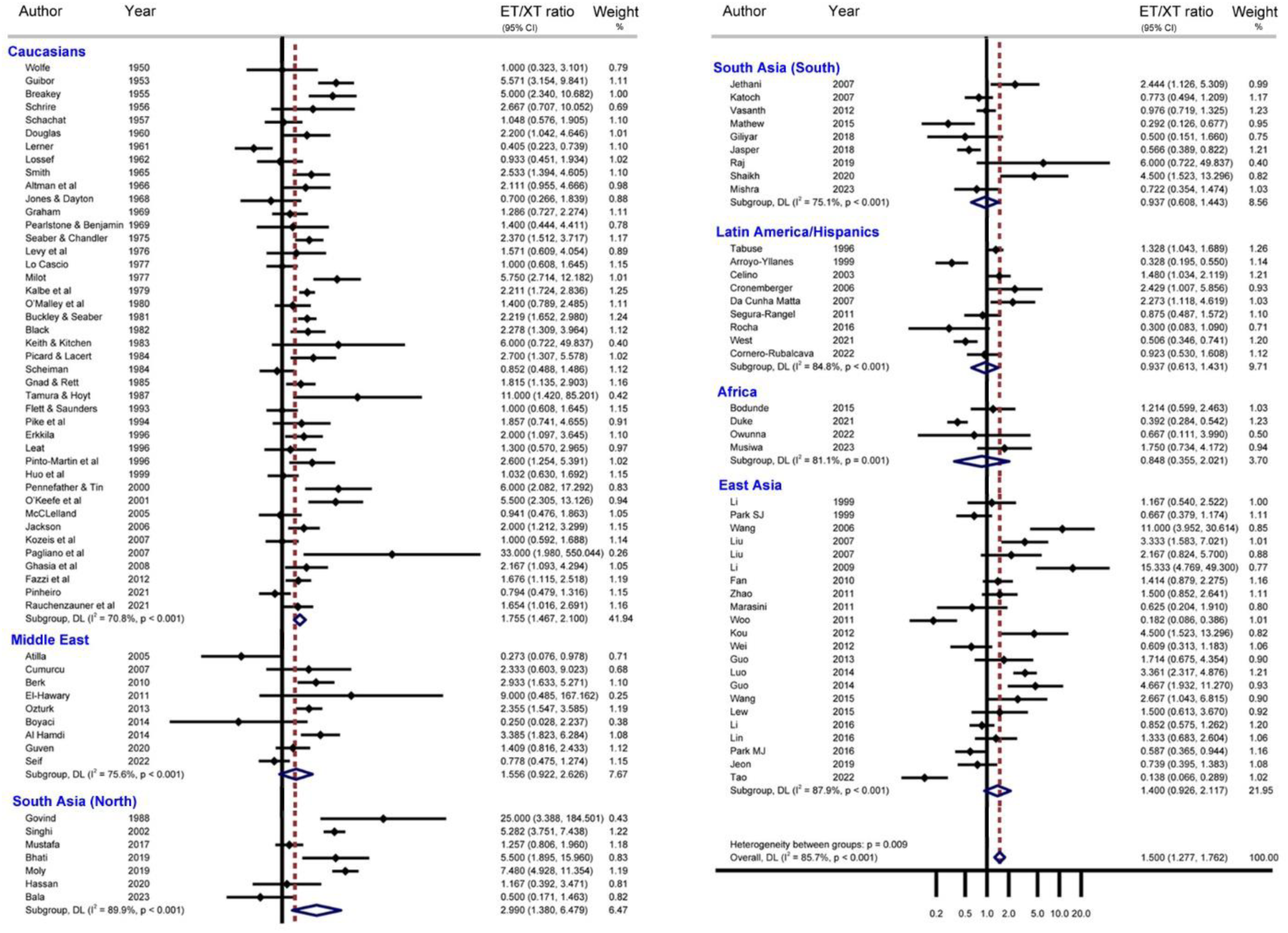
Forest Plot of Esotropia/Exotropia (ET/XT) Ratios in Cerebral Palsy sorted by Ethnicity. Ethnicities with higher ET/XT ratios (higher than 1.5) are compiled on the left side, ethnicities with lower ET/XT ratios (below 1.5, several below 1.0) are compiled on the right side of the forest plot. The diamonds show the pooled ratios for the listed studies.

**Fig. 5.**
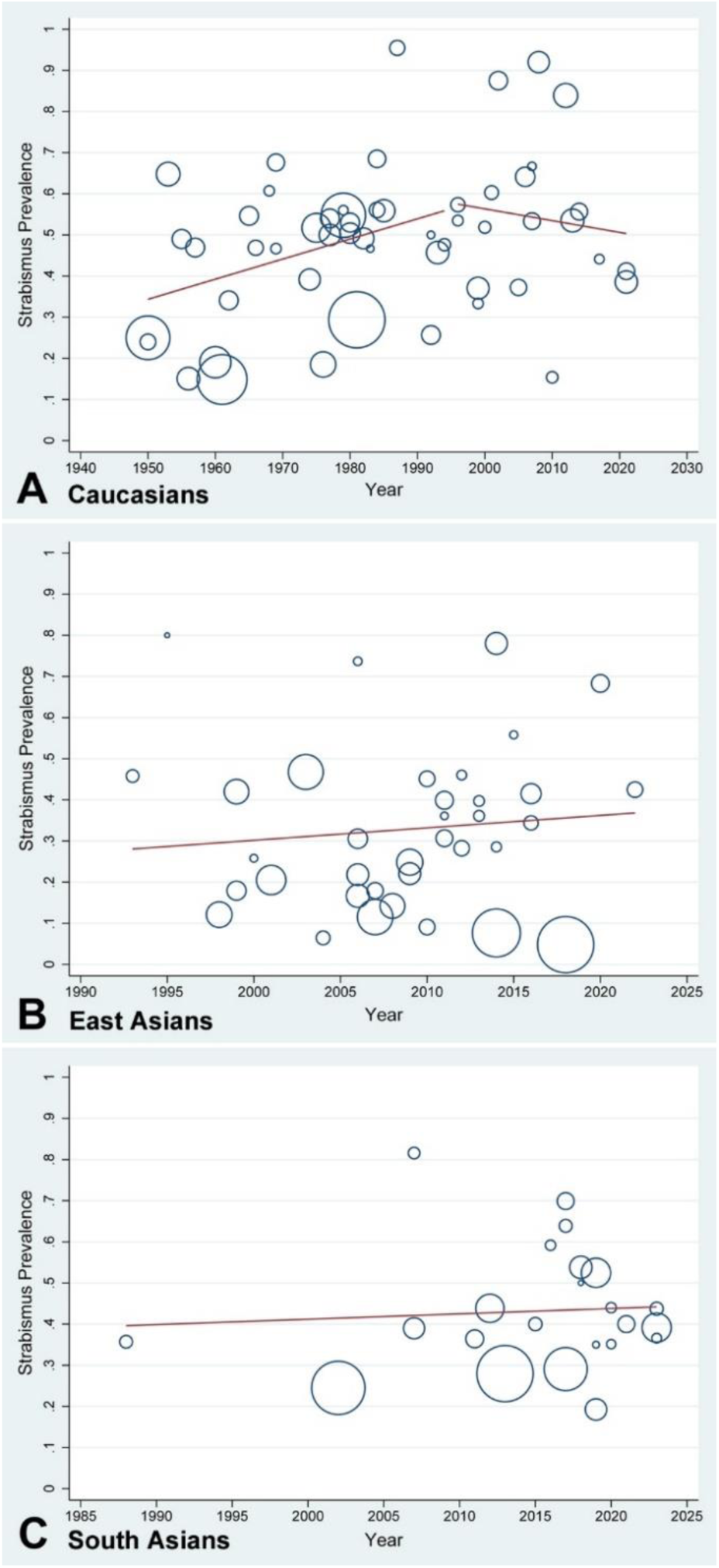
**A-C.** Trends over decades in the prevalence of strabismus in cerebral palsy (CP) in Caucasians (**A**), East Asians (**B**), and South Asians (**C**). Note the increasing trend from about 35% to 55% (significant with p=0.039) in Caucasians from 1950 to 1995, and a near stable trend thereafter (**A**). East Asians show a more gradual increase from 1995 to 2022 (**B**), while the trendline for South Asians is nearly flat (**C**). These data are consistent with the notion that with more effective maternal health care, the prevalence of strabismus in CP increases, likely due to fewer cases of CP with perinatal etiology and increased numbers of surviving premature babies, and then reaches a plateau.

### Gender distribution

In the CP cohorts examined for gender, males dominate over females by a ratio of 1.66:1. This ratio is similar to the known male dominance in CP (about 1.5 to 1).^184^ Among CP cases, the strabismus prevalence appears to be the same between males and females (although this is based on sparse data). Because of the male dominance in CP, there are more males with CP-associated strabismus than females. In the normal (non-CP) population, the prevalence of strabismus is the same between males and females.^206^ Our data suggest that there is a male predominance of CP-associated strabismus (7.6 million males vs 4.6 million females), which would indicate a 3 million male/female difference in the global cases of all strabismus, when CP-associated strabismus cases are included in the count.

### Pathogenesis of strabismus in CP

It is currently controversial what causes strabismus in CP; three different explanations have been proposed: (1) spasticity of extraocular muscle, (2) lesion of cortical or subcortical motor circuits, and (3) lesion of cortical or subcortical visual circuits. We will discuss these scenarios in sequence and evaluate their merits.

Extraocular muscle spasticity (similar to the limb spasticity seen in CP) as a mechanism for strabismus was discussed by Smith,^1,70^ and proposed by Sandfield-Nielsen et al.^200^ This notion was essentially refuted by Smith^1,70^ and Kalbe et al.^51^ It was pointed out that extraocular muscles are not spastic in CP, and there is no reason why any such “spasticity” should selectively affect certain extraocular muscles and spare others. While strabismic extraocular muscles do exhibit abnormal extracellular matrix gene and protein expression,^207^ which resembles abnormalities in CP limb skeletal muscle,^26^ such dysregulation may be a general response of muscle tissue to abnormal functional demands, and does not necessarily imply similarity in pathogenesis.

Some authors state that subcortical motor lesions (midbrain, brainstem) may cause strabismus in CP.^1,45,54,94,155,167^ Lesions of basal ganglia and cerebellum (dyskinetic and ataxic CP) less often associate with strabismus.^10,36,37,47,105,189^

Other authors implicate lesions of cortical or subcortical visual pathways as the primary cause of strabismus in CP.^1,46,56,56,189,208,209^ The significance of impaired visual pathways (optic radiations and/or cortical areas involved in visual motion processing) for strabismus in CP is supported by imaging studies (CT, MRI).^189,208,210–214^. Disturbance of motor pathways appears to play a lesser role in the pathogenesis of strabismus in CP than compromised visual pathways. It was noted that the lesion of subcortical visual pathways – more frequent in preterm births due to the immaturity of the pathways – appears to cause more esotropia, while lesions of visual cortex more often cause exotropia^189,214,215^ – but some authors disagree.^209^ Such differences in the direction of horizontal misalignment are consistent with prenatal lesions being more frequent in high-income countries, and perinatal lesions being more frequent in lower-income countries. Overall, the direction of horizontal strabismus in CP appears to be determined primarily by ethnic/genetic factors rather than socioeconomic factors or timing of lesions (prenatal vs perinatal). Regions with a lower ET/XT ratio (South India) utilize maternal health care more frequently than regions with a higher ET/XT ratio (North of India),^216^ so a shift to more prenatal insults than perinatal insults cannot explain the lower ET/XT ratio in South India (Fig. 2B).

### Conclusion

Differences in maternal health care between lower-income and high-income countries are important to understand global and regional differences in CP and to predict trends in CP prevalence as well as the prevalence of strabismus in CP.

## Data Availability

All data reviewed in this manuscript are available in the referenced sources.

## Acknowledgments

The authors thank Jenny Costa (Savitt Medical Library, University of Nevada, Reno) for assistance with finding sources. We also thank Dr. Mitchell Strominger (University of Nevada, Reno, School of Medicine) and Dr. Wei Yang (University of Nevada, Reno, School of Public Health) for helpful comments.

## SUPPLEMENTAL FILES

**Supplementary Table 1.**
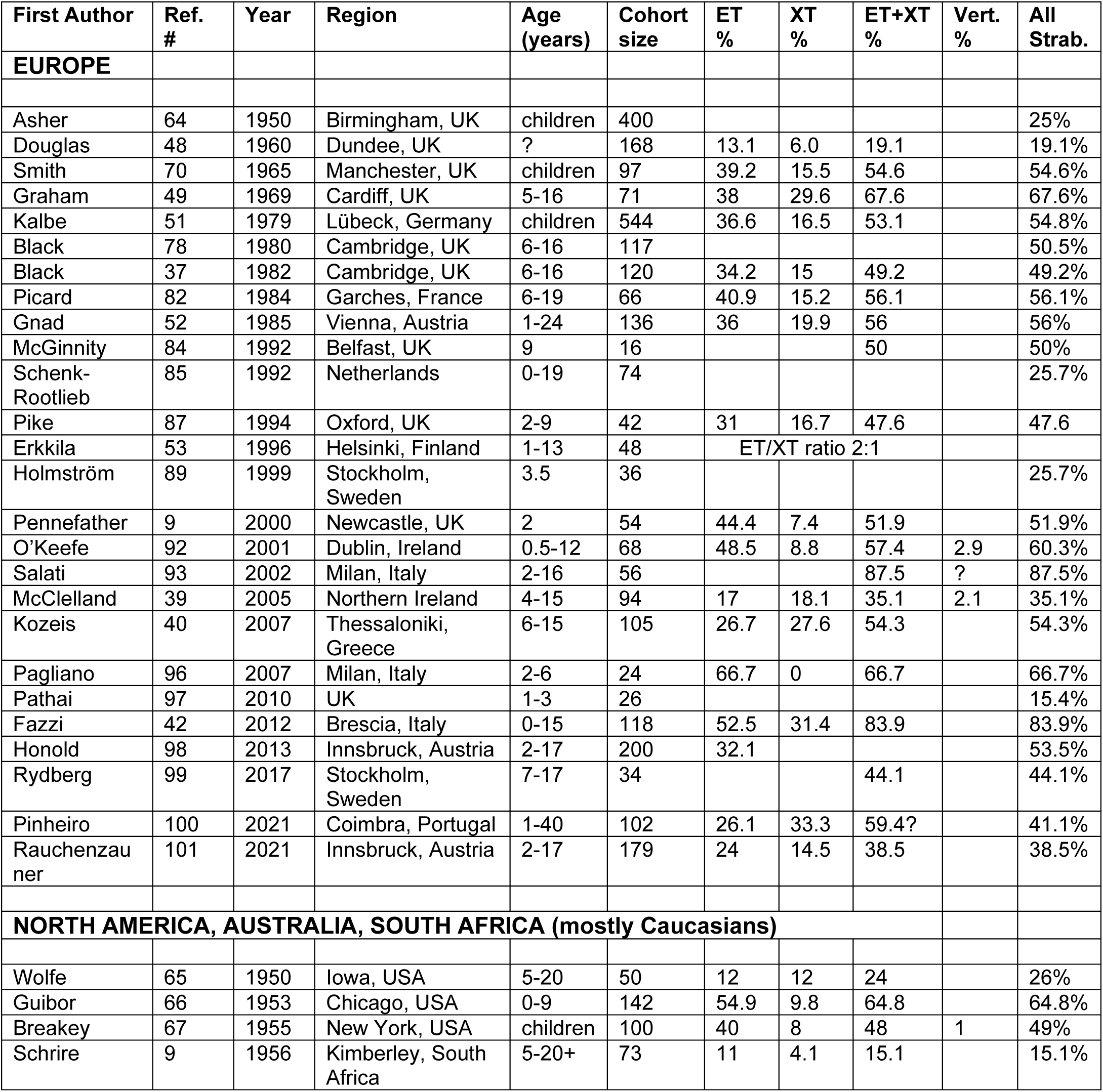
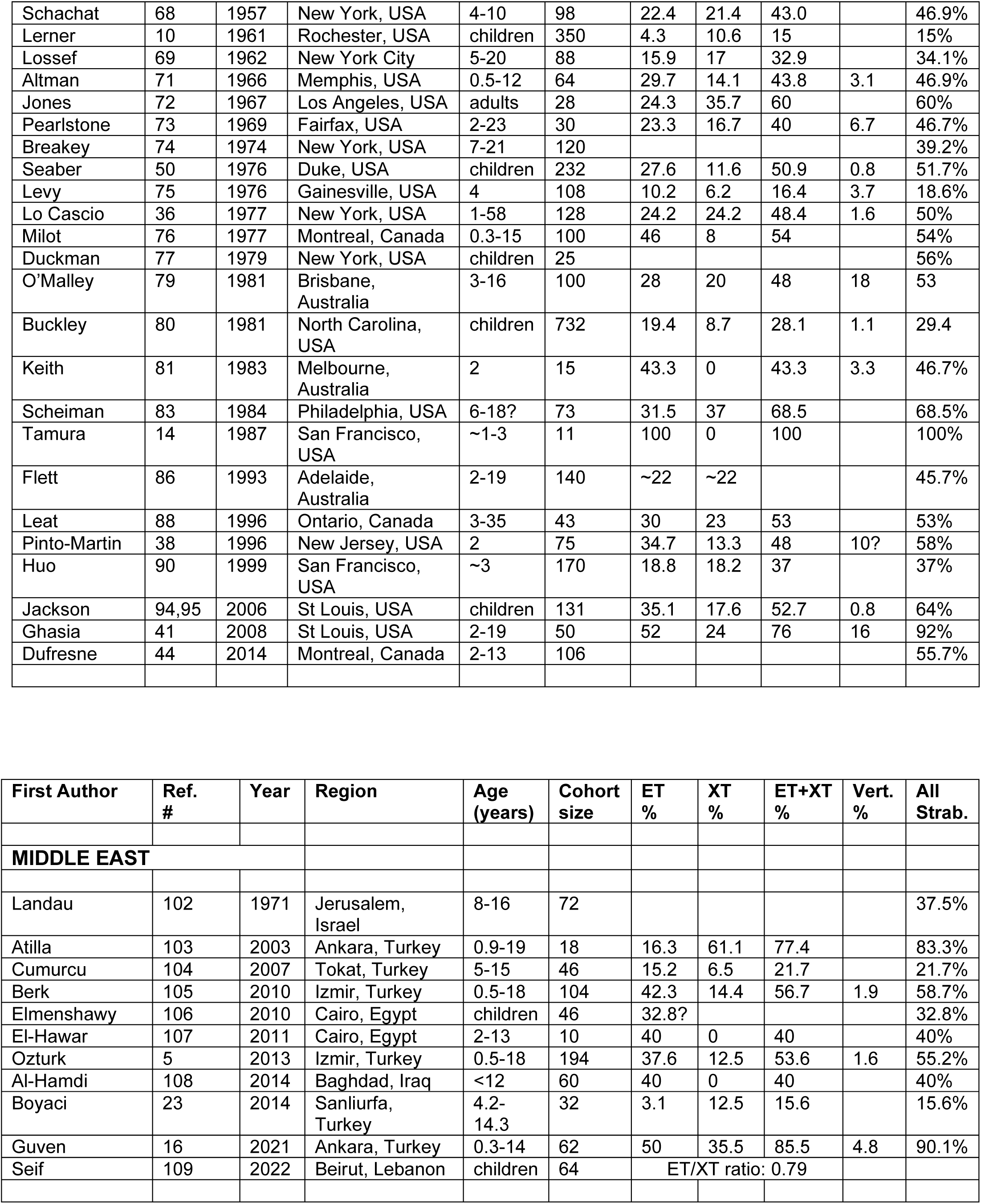
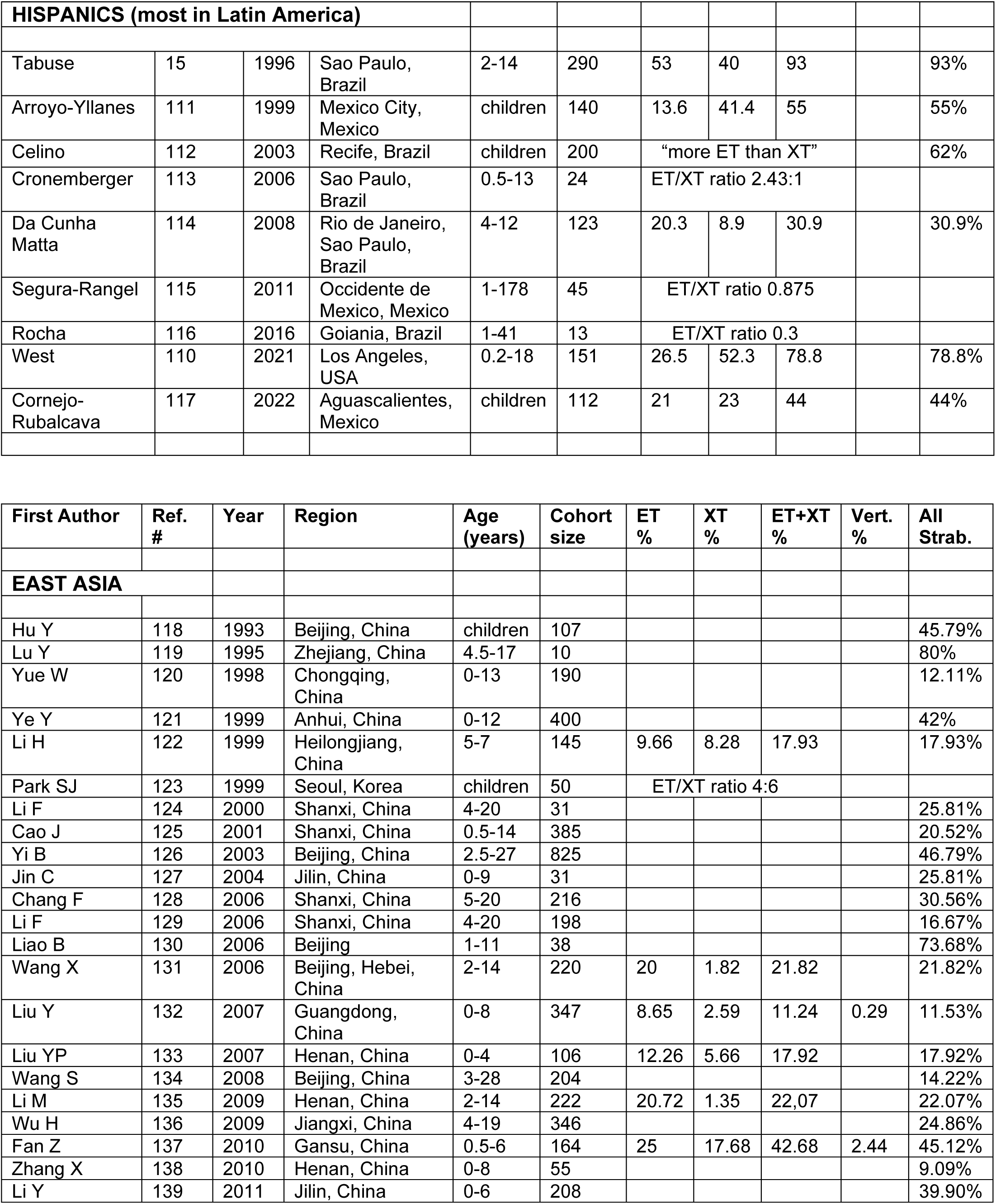
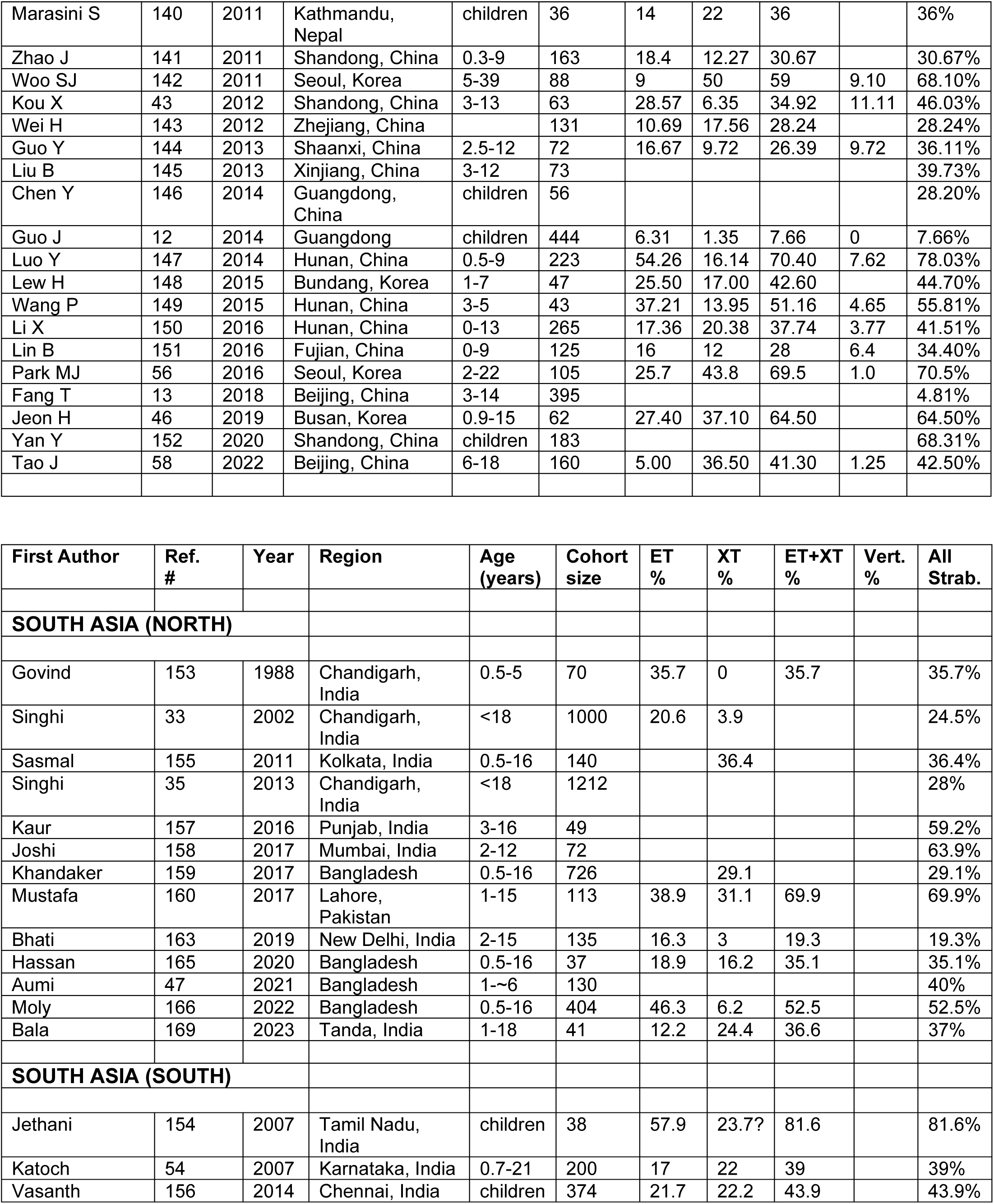
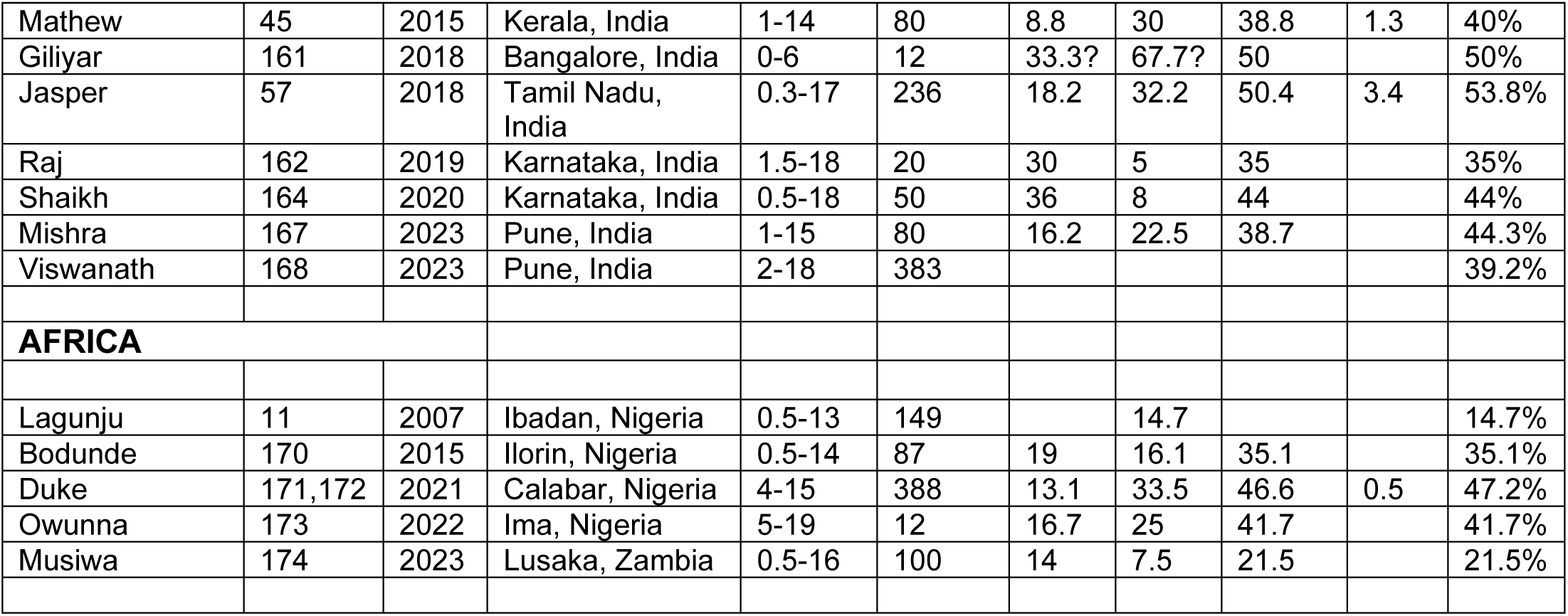
Prevalence of strabismus, esotropia (ET), exotropia (XT), vertical deviation (Vert.) and all strabismus (All Strab.) in cohort studies of cerebral palsy. Ref., Reference. The Table is organized by region/ethnicity as well as chronological within those regions. Explanations: ?, data are uncertain or inconsistent in the report; ∼, approximate numbers; +, a few subjects in the cohort exceeded the value; blank, no information was given. We list only one study in cases where multiple papers report on the same cohort.

**Supplemental Fig. 1.**
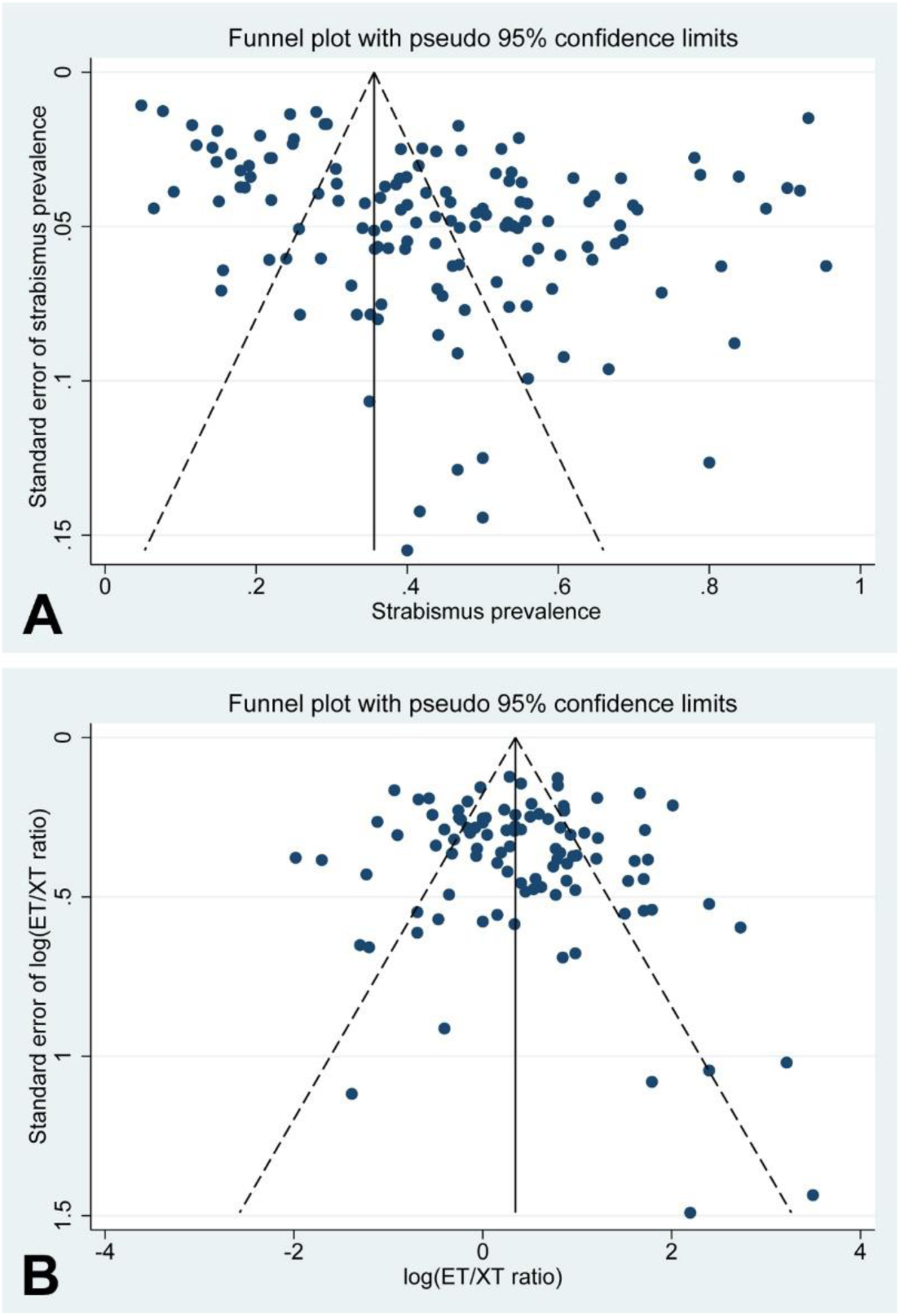
**A, B.** Funnel Plot for Prevalence Studies (**A**). Note that the asymmetry (p<0.001) arises because the central location of the funnel plot is at ∼35%. For studies with prevalence rates between 70% and 100% not to appear as outliers, the Funnel Plot would have to include prevalence data in a range of less than 0 to -30%, which is impossible for a prevalence. Esotropia/Exotropia (ET/XT) Ratio (**B**). There is no significant asymmetry in this Funnel Plot (p=0.381).

**Supplemental Fig. 2.**
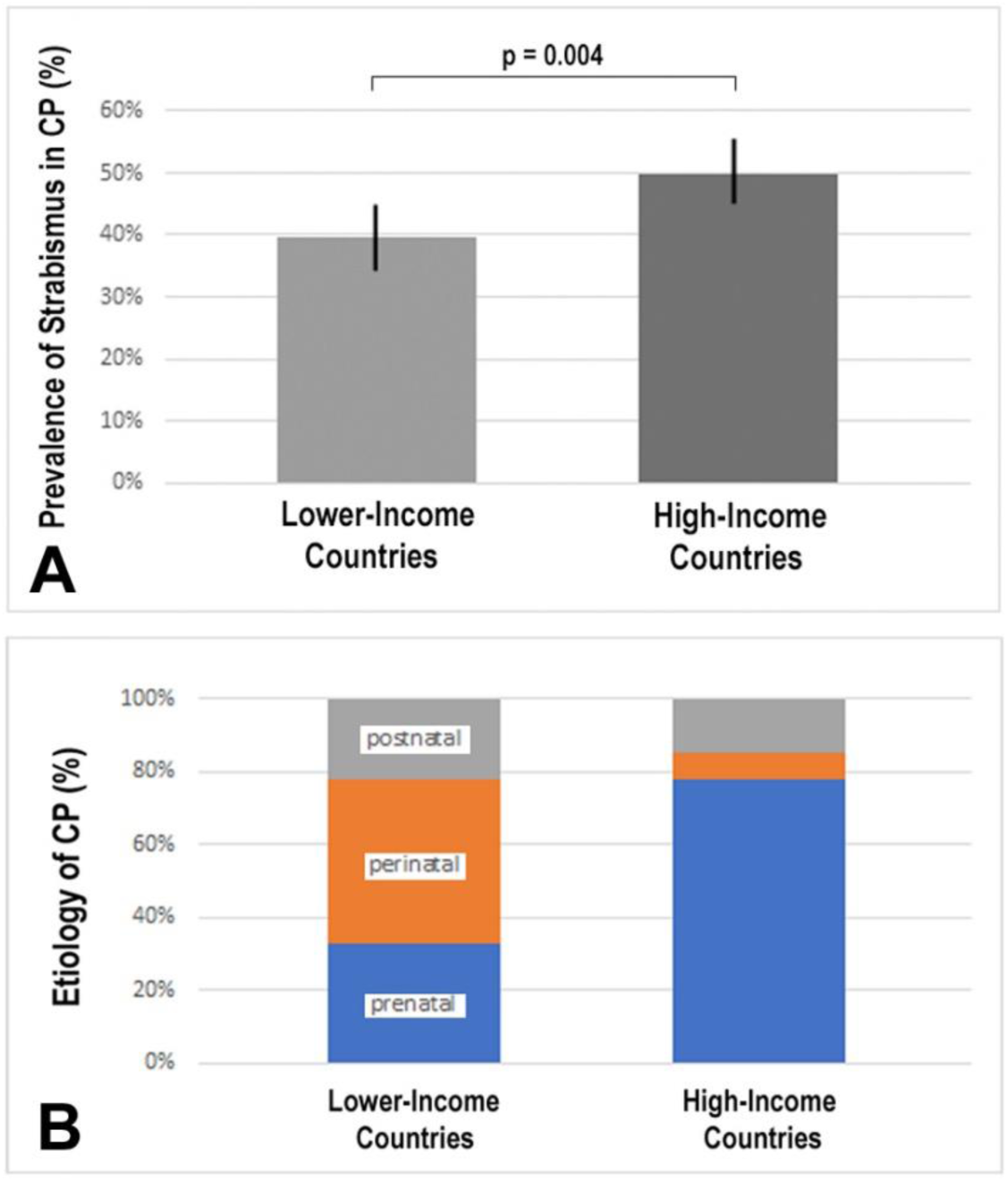
**A, B.** Graph illustrates the difference in strabismus prevalence in cerebral palsy (CP) between lower-income countries (LICs) and high-income countries (HICs) (**A**). Bars indicate the 95% confidence intervals; the difference is significant with p=0.004. Graph illustrates the differences in the etiology of CP between LICs and HICs: prenatal, perinatal and postnatal (**B**). The perinatal causes include mostly asphyxia, but also jaundice, seizures and infections. Prenatal causes include preterm births and low birthweight. CP etiology is represented in LICs according to Singhi et al.,^33^ and in HICs according to Reddihough and Collins.^217^

## Notes

**Funding Details:** This work was supported by the [National Institutes of Health] under Grant [EY031729]; [National Institutes of Health] under Grant [GM103554]; and [Office of Medical Research, University of Nevada, Reno, School of Medicine] under Grant [microgrant to MSH].

### Competing Interest Statement

The authors have declared no competing interest.

### Funding Statement

This work was supported by the National Institutes of Health under Grants EY031729 and GM103554, and a microgrant from the Office of Medical Research, University of Nevada, Reno, School of Medicine.

## REFERENCES

1. Smith V. A survey of strabismus in cerebral palsy. Little Club Clin Developmental Med. 1963;9:22–36.

2. Coppens A. L’aspect ophtalmologique de la paralysie cérébrale infantile ou infirmité mortrice cérébrale [Opthalmologic aspects of cerebral infantile palsy or cerebral motor infirmity]. Ann Ocul (Paris). 1967;200(2):191–217.

3. Hiles DA. Results of strabismus therapy in cerebral palsied children. Am Orthopt J. 1975;25:46–55.

4. Pigassou-Albouy R, Fleming A. Amblyopia and strabismus in patients with cerebral palsy. Ann Ophthalmol. 1975;7(3):382–384, 386-387.

5. Ozturk AT, Berk AT, Yaman A. Ocular disorders in children with spastic subtype of cerebral palsy. Int J Ophthalmol. 2013;6(2):204–210. doi:10.3980/j.issn.2222-3959.2013.02.19

6. Heydarian S, Abbasabadi MM, Khabazkhoob M, Hoseini-Yazdi H, Gharib M. Vision Abnormalities in Children and Young Adults With Cerebral Palsy; A Systematic Review. Semin Ophthalmol. 2022;37(4):471–479. doi:10.1080/08820538.2021.2021248

7. Hashemi H, Pakzad R, Heydarian S, et al. Global and regional prevalence of strabismus: a comprehensive systematic review and meta-analysis. Strabismus. 2019;27(2):54–65. doi:10.1080/09273972.2019.1604773

8. Adinanto FC. Development of Vision and Strabismus in Childhood: Prevalence and Risk Factors. Thesis. 2020. Accessed September 27, 2023. https://opus.lib.uts.edu.au/handle/10453/147289

9. Schrire L. An ophthalmological survey of a series of cerebral palsy cases. S Afr Med J. 1956;30(17):405–407.

10. Lerner HA. Spastic and cerebral palsy squint problems. Am Orthopt J. 1961;11:72–75.

11. Lagunju IA, Oluleye TS. Ocular abnormalities in children with cerebral palsy. Afr J Med Med Sci. 2007;36(1):71–75.

12. Guo J. Clinical Study on Comorbidities in Children with Cerebral Palsy. Master’s thesis, Guangzhou University of Chinese Medicine. Published online 2014.

13. Fang T, Xu J, Xie Z. The clinical study of selective posterior rhizotomy and vertebral reduction for the treatment of spastic cerebral palsy. Chin J Neurosurg Dis Res. 2018;(06):532–536.

14. Tamura EE, Hoyt CS. Oculomotor consequences of intraventricular hemorrhages in premature infants. Arch Ophthalmol. 1987;105(4):533–535. doi:10.1001/archopht.1987.01060040103043

15. Tabuse MKU, Moreira JB de C. Ocular abnormalities in children with cerebral palsy. Arq Bras Oftalmol. 1996;59:560–567. doi:10.5935/0004-2749.19960004

16. Guven S, Uludag HA, Kucukevcilioglu M, Mutlu FM. Ocular characteristics of patients with cerebral palsy. Ann Med Res. 2020;27(12):3112–3116.

17. Stanley F, Blair E, Alberman E. Cerebral Palsies: Epidemiology and Causal Pathways. Mac Keith Press; 2000.

18. Odding E, Roebroeck ME, Stam HJ. The epidemiology of cerebral palsy: incidence, impairments and risk factors. Disabil Rehabil. 2006;28(4):183–191. doi:10.1080/09638280500158422

19. Oskoui M, Coutinho F, Dykeman J, Jetté N, Pringsheim T. An update on the prevalence of cerebral palsy: a systematic review and meta-analysis. Dev Med Child Neurol. 2013;55(6):509–519. doi:10.1111/dmcn.12080

20. McIntyre S, Goldsmith S, Webb A, et al. Global prevalence of cerebral palsy: A systematic analysis. Dev Med Child Neurol. 2022;64(12):1494–1506. doi:10.1111/dmcn.15346

21. Couper J. Prevalence of childhood disability in rural KwaZulu-Natal. S Afr Med J. 2002;92(7):549–552.

22. Serdaroğlu A, Cansu A, Ozkan S, Tezcan S. Prevalence of cerebral palsy in Turkish children between the ages of 2 and 16 years. Dev Med Child Neurol. 2006;48(6):413–416. doi:10.1017/S0012162206000910

23. Boyaci A, Akal A, Tutoglu A, et al. Relationship among Ocular Diseases, Developmental Levels, and Clinical Characteristics of Children with Diplegic Cerebral Palsy. J Phys Ther Sci. 2014;26(11):1679–1684. doi:10.1589/jpts.26.1679

24. Colver A, Fairhurst C, Pharoah POD. Cerebral palsy. Lancet. 2014;383(9924):1240–1249. doi:10.1016/S0140-6736(13)61835-8

25. Donald KA, Samia P, Kakooza-Mwesige A, Bearden D. Pediatric cerebral palsy in Africa: a systematic review. Semin Pediatr Neurol. 2014;21(1):30–35. doi:10.1016/j.spen.2014.01.001

26. Graham HK, Rosenbaum P, Paneth N, et al. Cerebral palsy. Nat Rev Dis Primers. 2016;2:15082. doi:10.1038/nrdp.2015.82

27. Kakooza-Mwesige A, Andrews C, Peterson S, Wabwire Mangen F, Eliasson AC, Forssberg H. Prevalence of cerebral palsy in Uganda: a population-based study. Lancet Glob Health. 2017;5(12):e1275–e1282. doi:10.1016/S2214-109X(17)30374-1

28. Khandaker G, Muhit M, Karim T, et al. Epidemiology of cerebral palsy in Bangladesh: a population-based surveillance study. Dev Med Child Neurol. 2019;61(5):601–609. doi:10.1111/dmcn.14013

29. Cieza A, Causey K, Kamenov K, Hanson SW, Chatterji S, Vos T. Global estimates of the need for rehabilitation based on the Global Burden of Disease study 2019: a systematic analysis for the Global Burden of Disease Study 2019. Lancet. 2021;396(10267):2006–2017. doi:10.1016/S0140-6736(20)32340-0

30. Jahan I, Muhit M, Hardianto D, et al. Epidemiology of cerebral palsy in low- and middle-income countries: preliminary findings from an international multi-centre cerebral palsy register. Dev Med Child Neurol. 2021;63(11):1327–1336. doi:10.1111/dmcn.14926

31. Olusanya BO, Gladstone M, Wright SM, et al. Cerebral palsy and developmental intellectual disability in children younger than 5 years: Findings from the GBD-WHO Rehabilitation Database 2019. Frontiers in Public Health. 2022;10. Accessed December 21, 2023. https://www.frontiersin.org/articles/10.3389/fpubh.2022.894546

32. Barron-Garza F, Coronado-Garza M, Gutierrez-Ramirez S, et al. Incidence of Cerebral Palsy, Risk Factors, and Neuroimaging in Northeast Mexico. Pediatr Neurol. 2023;143:50–58. doi:10.1016/j.pediatrneurol.2023.02.005

33. Singhi PD, Ray M, Suri G. Clinical Spectrum of Cerebral Palsy in North India—An Analysis of 1000 Cases. Journal of Tropical Pediatrics. 2002;48(3):162–166. doi:10.1093/tropej/48.3.162

34. Gladstone M. A review of the incidence and prevalence, types and aetiology of childhood cerebral palsy in resource-poor settings. Ann Trop Paediatr. 2010;30(3):181–196. doi:10.1179/146532810X12786388978481

35. Singhi P, Saini AG. Changes in the clinical spectrum of cerebral palsy over two decades in North India--an analysis of 1212 cases. J Trop Pediatr. 2013;59(6):434–440. doi:10.1093/tropej/fmt035

36. Lo Cascio GP. A study of vision in cerebral palsy. Am J Optom Physiol Opt. 1977;54(5):332–337. doi:10.1097/00006324-197705000-00011

37. Black P. Visual disorders associated with cerebral palsy. Br J Ophthalmol. 1982;66(1):46–52. doi:10.1136/bjo.66.1.46

38. Pinto-Martin JA, Dobson V, Cnaan A, Zhao H, Paneth NS. Vision outcome at age 2 years in a low birth weight population. Pediatr Neurol. 1996;14(4):281–287. doi:10.1016/0887-8994(96)00051-3

39. McClelland JF, Jackson AJ, Parkes J, Hill N, Saunders KJ. Visual Acuity and Disorders of Ocular Posture in Children With Cerebral Palsy. Investigative Ophthalmology & Visual Science. 2005;46(13):1935.

40. Kozeis N, Anogeianaki A, Mitova DT, Anogianakis G, Mitov T, Klisarova A. Visual function and visual perception in cerebral palsied children. Ophthalmic Physiol Opt. 2007;27(1):44–53. doi:10.1111/j.1475-1313.2006.00413.x

41. Ghasia F, Brunstrom J, Gordon M, Tychsen L. Frequency and Severity of Visual Sensory and Motor Deficits in Children with Cerebral Palsy: Gross Motor Function Classification Scale. Investigative Ophthalmology & Visual Science. 2008;49(2):572–580. doi:10.1167/iovs.07-0525

42. Fazzi E, Signorini SG, LA Piana R, et al. Neuro-ophthalmological disorders in cerebral palsy: ophthalmological, oculomotor, and visual aspects. Dev Med Child Neurol. 2012;54(8):730–736. doi:10.1111/j.1469-8749.2012.04324.x

43. Kou X, Kong Q, Ni L, Yin X. CHILDREN WITH SPASTIC CEREBRAL PALSY: A GAZE-POSITION ANALYSIS. Med J Qilu. 2012;(02):158–160.

44. Dufresne D, Dagenais L, Shevell MI, REPACQ Consortium. Spectrum of visual disorders in a population-based cerebral palsy cohort. Pediatr Neurol. 2014;50(4):324–328. doi:10.1016/j.pediatrneurol.2013.11.022

45. Mathew E, Jacob JM, Goudinho S. Prevalence of Strabismus in Patients with Cerebral Palsy. Int J Advances Health Sci. 2015;2:120–128.

46. Jeon H, Jung JH, Yoon JA, Choi H. Strabismus Is Correlated with Gross Motor Function in Children with Spastic Cerebral Palsy. Curr Eye Res. 2019;44(11):1258–1263. doi:10.1080/02713683.2019.1631851

47. Aumi W, Afroz F, Maksud S. Ocular defect in children with cerebral palsy and its correlation with the types of cerebral palsy. Int J Contemp Pediatr. 2021;8:609–615.

48. Douglas A. The eyes and vision in infantile cerebral palsy. Trans Ophthalmol Soc UK. 1960;80:311–325.

49. Graham MV. The spastic child. Proc R Soc Med. 1969;62(6):563–564.

50. Seaber J, Chandler A. A five-year study of patients with cerebral palsy and strabismus: orthoptics: past, present, future. In: Transactions of Third International Orthoptic Congress. Stratton Intercontinental Medical Book Corp.; 1976:271-277.

51. Kalbe U, Berndt K, de Decker W. Strabismus bei zerebralparetischen und ungeschädigten Kindern. Vergleich der motorischen Symptome [Strabismus in cerebral paretic and normal children. Comparison of motoric symptoms (author’s transl)]. Klin Monbl Augenheilkd. 1979;175(3):367–374.

52. Gnad H, Rett A. [Ophthalmological symptoms of infantile cerebral palsy]. Wien Klin Wochenschr. 1985;97(19):749–752.

53. Erkkilä H, Lindberg L, Kallio AK. Strabismus in children with cerebral palsy. Acta Ophthalmol Scand. 1996;74(6):636–638. doi:10.1111/j.1600-0420.1996.tb00752.x

54. Katoch S, Devi A, Kulkarni P. Ocular defects in cerebral palsy. Indian J Ophthalmol. 2007;55(2):154–156. doi:10.4103/0301-4738.30717

55. Ma DJ, Yang HK, Hwang JM. Surgical responses and outcomes of bilateral lateral rectus recession in exotropia with cerebral palsy. Acta Ophthalmol. 2017;95(3):e179–e184. doi:10.1111/aos.13158

56. Park MJ, Yoo YJ, Chung CY, Hwang JM. Ocular findings in patients with spastic type cerebral palsy. BMC Ophthalmol. 2016;16:195. doi:10.1186/s12886-016-0367-1

57. Jasper S, Philip SS. Profile of Cerebral Visual Impairment in Children with Cerebral Palsy at a Tertiary Care Referral Center in Southern India. JCDR. Published online 2018. doi:10.7860/JCDR/2018/33984.11316

58. Tao J, Hao R, Guo Y, Zhang W. Characteristics of Visual Function in Children with Cerebral Palsy and Mental Retardation in Urban Beijing. Research Square [Preprint].; 2022. doi:10.21203/rs.3.rs-1827694/v1

59. Ye H, Liu Q, Zhan Q, et al. Surgical outcomes and observation in exotropia cerebral palsy children with cortical visual impairment. BMC Ophthalmol. 2022;22(1):364. doi:10.1186/s12886-022-02581-x

60. Herron MS, Wang L, von Bartheld CS. Prevalence and types of strabismus in cerebral palsy: A global and historical perspective based on a systematic review and meta-analysis. [Preprint]. Published online 2024.

61. Moher D, Liberati A, Tetzlaff J, Altman DG, PRISMA Group. Preferred reporting items for systematic reviews and meta-analyses: the PRISMA statement. PLoS Med. 2009;6(7):e1000097. doi:10.1371/journal.pmed.1000097

62. Rosenbaum P, Paneth N, Leviton A, et al. A report: the definition and classification of cerebral palsy April 2006. Dev Med Child Neurol Suppl. 2007;109:8–14.

63. Alimovic S. Visual Impairments in Children with Cerebral Palsy. Hrvatska Revija Za Rehabilitacijska Istrazivanja. 2012;48.

64. Asher P, Schonell FE. A survey of 400 cases of cerebral palsy in childhood. Arch Dis Child. 1950;25(124):360–379. doi:10.1136/adc.25.124.360

65. Wolfe WG. A comprehensive evaluation of fifty cases of cerebral palsy. J Speech Disord. 1950;15(3):234–251. doi:10.1044/jshd.1503.234

66. Guibor GP. Some eye defects seen in cerebral palsy, with some statistics. Am J Phys Med. 1953;32(6):342–347.

67. Breakey AS. Ocular Findings in Cerebral Palsy. AMA Archives of Ophthalmology. 1955;53(6):852–856. doi:10.1001/archopht.1955.00930010860011

68. Schachat WS, Wallace HM, Palmer M, Slater B. Ophthalmologic findings in children with cerebral palsy. Pediatrics. 1957;19(4 Pt 1):623–628.

69. Lossef S. Ocular Findings in Cerebral Palsy* *From the Bureau for Handicapped Children, New York City Department of Health, and the Bird S. Coler Hospital Children’s Rehabilitation, New York Medical College. American Journal of Ophthalmology. 1962;54(6):1114–1118. doi:10.1016/0002-9394(62)94353-2

70. Smith V. Strabismus in cerebral palsy. Br Orthopt J. 1965;22:84–94.

71. Altman HE, Hiatt RL, Deweese MW. Ocular findings in cerebral palsy. South Med J. 1966;59(9):1015–1018. doi:10.1097/00007611-196609000-00005

72. Jones M, Dayton GO. Assessment of visual disorders in cerebral palsy. Archivio italiano di pediatria e puericoltura. Published online 1967. Accessed September 27, 2023. https://www.semanticscholar.org/paper/Assessment-of-visual-disorders-in-cerebral-palsy.-Jones-Dayton/554555a2028bb1fe4ad992322d01591913337b9e

73. Pearlstone A, Benjamin R. Ocular defects in cerebral palsy. Eye Ear Nose Throat Mon. 1969;48(7):437–439.

74. Breakey AS, Wilson JJ, Wilson BC. Sensory and perceptual functions in the cerebral palsied. 3. Some visual perceptual relationships. J Nerv Ment Dis. 1974;158(1):70–77. doi:10.1097/00005053-197401000-00009

75. Levy NS, Cassin B, Newman M. Strabismus in children with cerebral palsy. J Pediatr Ophthalmol. 1976;13(2):72–74.

76. Milot J, Guimond J. Strabismus as an expression of cerebral motor dysfunction in childhood. Clinical comments based on 54 instances among 100 such children in Montreal. Clin Pediatr (Phila). 1977;16(5):477–479. doi:10.1177/000992287701600514

77. Duckman R. The incidence of visual anomalies in a population of cerebral palsied children. J Am Optom Assoc. 1979;50(9):1013–1016.

78. Black PD. Ocular defects in children with cerebral palsy. Br Med J. 1980;281(6238):487–488. doi:10.1136/bmj.281.6238.487

79. O’Malley J, Stark DJ, Manning L, Cowley H. An Ophthalmic Review of Cerebral Palsy in Queensland 1980. Australian Journal of Opthalmology. 1981;9(2):91–95. doi:10.1111/j.1442-9071.1981.tb01495.x

80. Buckley E, Seaber JH. Dyskinetic strabismus as a sign of cerebral palsy. Am J Ophthalmol. 1981;91(5):652–657. doi:10.1016/0002-9394(81)90069-6

81. Keith CG, Kitchen WH. Ocular morbidity in infants of very low birth weight. Br J Ophthalmol. 1983;67(5):302–305. doi:10.1136/bjo.67.5.302

82. Picard A, Lacert P. Les troubles de la motricité horizontale du regard chez l’infirme moteur cérébral [Disorders of horizontal gaze motility in the cerebral palsy patient]. J Fr Ophtalmol. 1984;7(11):717–720.

83. Scheiman MM. Optometric findings in children with cerebral palsy. Am J Optom Physiol Opt. 1984;61(5):321–323. doi:10.1097/00006324-198405000-00005

84. McGinnity FG, Bryars JH. Controlled study of ocular morbidity in school children born preterm. British Journal of Ophthalmology. 1992;76(9):520–524. doi:10.1136/bjo.76.9.520

85. Schenk-Rootlieb AJ, van Nieuwenhuizen O, van der Graaf Y, Wittebol-Post D, Willemse J. The prevalence of cerebral visual disturbance in children with cerebral palsy. Dev Med Child Neurol. 1992;34(6):473–480. doi:10.1111/j.1469-8749.1992.tb11467.x

86. Flett P, Saunders B. Ophthalmic assessment of physically disabled children attending a rehabilitation centre. J Paediatr Child Health. 1993;29(2):132–135. doi:10.1111/j.1440-1754.1993.tb00465.x

87. Pike MG, Holmstrom G, de Vries LS, et al. Patterns of visual impairment associated with lesions of the preterm infant brain. Dev Med Child Neurol. 1994;36(10):849–862. doi:10.1111/j.1469-8749.1994.tb11776.x

88. Leat SJ. Reduced accommodation in children with cerebral palsy. Ophthalmic Physiol Opt. 1996;16(5):385–390.

89. Holmstrom G, el Azazi M, Kugelberg U. Ophthalmological follow up of preterm infants: a population based, prospective study of visual acuity and strabismus. Br J Ophthalmol. 1999;83(2):143–150.

90. Huo R, Burden SK, Hoyt CS, Good WV. Chronic cortical visual impairment in children: aetiology, prognosis, and associated neurological deficits. Br J Ophthalmol. 1999;83(6):670–675. doi:10.1136/bjo.83.6.670

91. Pennefather PM, Tin W. Ocular abnormalities associated with cerebral palsy after preterm birth. Eye (Lond). 2000;14 ( Pt 1):78–81. doi:10.1038/eye.2000.17

92. O’Keefe M, Kafil-Hussain N, Flitcroft I, Lanigan B. Ocular significance of intraventricular haemorrhage in premature infants. Br J Ophthalmol. 2001;85(3):357–359. doi:10.1136/bjo.85.3.357

93. Salati R, Borgatti R, Giammari G, Jacobson L. Oculomotor dysfunction in cerebral visual impairment following perinatal hypoxia. Dev Med Child Neurol. 2002;44(8):542–550. doi:10.1017/s0012162201002535

94. Jackson J, Castleberry C, Galli M, Arnoldi KA. Cerebral Palsy for the Pediatric Eye Care Team Part II: Diagnosis and Treatment of Ocular Motor Deficits. Am Orthopt J. 2006;56:86–96. doi:10.3368/aoj.56.1.86

95. Arnoldi KA, Pendarvis L, Jackson J, Batra NNA. Cerebral Palsy for the Pediatric Eye Care Team Part III: Diagnosis and Management of Associated Visual and Sensory Disorders. Am Orthopt J. 2006;56:97–107. doi:10.3368/aoj.56.1.97

96. Pagliano E, Fedrizzi E, Erbetta A, et al. Cognitive profiles and visuoperceptual abilities in preterm and term spastic diplegic children with periventricular leukomalacia. J Child Neurol. 2007;22(3):282–288. doi:10.1177/0883073807300529

97. Pathai S, Cumberland PM, Rahi JS. Prevalence of and early-life influences on childhood strabismus: findings from the Millennium Cohort Study. Arch Pediatr Adolesc Med. 2010;164(3):250–257. doi:10.1001/archpediatrics.2009.297

98. Honold M, Baldissera I, Gedik A, et al. Visual disorders in children with cerebral palsy. Neuropediatrics. 2013;44(2):PS11_1027. doi:10.1055/s-0033-1337756

99. Rydberg A, Ygge J, Olsson M. Ocular Motor Function in Children with Spastic Hemiplegia Evaluated by the Ocular Motor Score. Strabismus. 2017;25(3):156–159. doi:10.1080/09273972.2017.1350727

100. Pinheiro RL, Cortez L, Paiva C, Murta JN. Characteristics of Patients Attending an Ophthalmic Outpatient Department at the Cerebral Palsy Association of Coimbra. Revista Sociedade Portuguesa de Oftalmologia. 2021;45(2):71–77. doi:10.48560/rspo.23784

101. Rauchenzauner M, Schiller K, Honold M, et al. Visual Impairment and Functional Classification in Children with Cerebral Palsy. Neuropediatrics. 2021;52(5):383–389. doi:10.1055/s-0040-1722679

102. Landau L, Berson D. Cerebral Palsy and Mental Retardation : Ocular Findings. Journal of Pediatric Ophthalmology & Strabismus. 1971;8(4):245–248. doi:10.3928/0191-3913-19711101-08

103. Atilla H, Batioğlu F, Ertuğrul C, Necile E. Ocular abnormalities associated with cerebral palsy. In: 29th European Strabismological Association Meeting, Izmir. CRC Press; 2006.

104. Cumurcu T, Cumurcu H, Erkorkmaz Ü, Yardim H. [Ocular findings in children with cerebral palsy.]. Firat J Med. 2007;12:48–52.

105. Berk T, Ozturk T, Yaman A. Ocular Disorders in Children with Cerebral Palsy. Türk Oftalmoloji Dergisi. 2010;40:209–216. doi:10.4274/tod.40.209

106. Elmenshawy AA, Ismael A, Elbehairy H, Kalifa NM, Fathy MA, Ahmed, Azzaw M. Visual Impairment in Children with Cerebral Palsy. International Journal of Academic Research. 2010;2(5):67–71.

107. El-Hawary GR, Shawky RM, El-Din AS, El-Din SMN. Ocular features in Egyptian genetically disabled children. Egyptian Journal of Medical Human Genetics. 2011;12(2):171–181. doi:10.1016/j.ejmhg.2011.06.004

108. Al-Hamdi A, Mahmood A, Kareem S. Assessment of Strabismus in Patients with Cerebral Palsy. Medical Journal of Babylon. 2014;11(1).

109. Seif R, Hmaimess G, Eid H, Dunya I. Strabismus Repair in Children with Varying Severity of Cerebral Palsy. Semin Ophthalmol. 2022;37(2):265–267. doi:10.1080/08820538.2021.2003823

110. West MR, Borchert MS, Chang MY. Ophthalmologic characteristics and outcomes of children with cortical visual impairment and cerebral palsy. Journal of American Association for Pediatric Ophthalmology and Strabismus. 2021;25(4):223.e1–223.e6. doi:10.1016/j.jaapos.2021.03.011

111. Arroyo-Yllanes ME, Manzo-Villalobos G, Perez-Perez JF, Garrido E. Strabismus in Patients with Cerebral Palsy. American Orthoptic Journal. 1999;49(1):141–147. doi:10.1080/0065955X.1999.11982204

112. Celino AC, Trigueiro S, Ventura LO, Toscano J, Barroca R. Alterações oculares em crianças portadoras de paralisia cerebral. Rev bras oftalmol. Published online 2003:248–251.

113. Cronemberger M, Mendonça T, Bicas H. Toxina botulínica no tratamento de estrabismo horizontal em crianças com paralisia cerebral [Botulinum toxin treatment for horizontal strabismus in children with cerebral palsy]. Arg Bras Oftalmol. 2006;69(4):523–529. doi:10.1590/s0004-27492006000400013

114. da Cunha Matta AP, Nunes G, Rossi L, Lawisch V, Dellatolas G, Braga L. Outpatient evaluation of vision and ocular motricity in 123 children with cerebral palsy. Developmental Neurorehabilitation. 2008;11(2):159–165. doi:10.1080/17518420701783622

115. Segura-Rangel I, Castellanos-Valencia A. Aplicación de toxina botulínica en niños con estrabismo y parálisis cerebral en un centro de rehabilitación. Rev Mex Oftalmol. 2011;85(4):189–195.

116. Rocha MNAM, Sanches A, Pessoa FF, et al. Clinical forms and risk factors associated with strabismus in visual binocularity. Rev bras.oftalmol. 2016;75:34–39. doi:10.5935/0034-7280.20160008

117. Cornejo-Rubalcava MF, Zamilpa-Velázquez F de R, Muñoz-Guerrero D, Casillas-Casillas E. Estado visual en niños con parálisis cerebral en el Centro de Rehabilitación e Inclusión Infantil Teletón Aguascalientes. Lux Médica. 2022;17(49). doi:10.33064/49lm20223274

118. Hu Y. Clinical Analysis of Ocular Functional Disorders in Children with Cerebral Palsy. J Appl Clin Pediatr. 1993;(Z1):377–378.

119. Lu Y, Huang H, Yang Y, Wang Y. Selective Dorsal Rhizotomy for Treating Spastic Cerebral Palsy. Mil Med J Southea Chin. 1995;(05):9–10.

120. Yue W, Zhang L, Duan J, Ma J. Analysis of CT Changes and Their Clinical Correlations in 190 Cases of Pediatric Cerebral Palsy. Sichuan MedJ. 1998;(05).

121. Ye Y. Discussion on the Incidence, Types, and Prognosis of Strabismus in Children with Cerebral Palsy. J Clin Ophthalmol. 1999;(04).

122. Li H, Li X, Zhang L, Wang L, Zhang L, Liu G. Survey on amblyopia in cerebral palsy children. Mod Rehabil. 1999;(12):1416–1417.

123. Park SJ, Chang BL. Stabismus, Amblyopia and Refractive Errors in Patients with Cerebral Palsy. Journal of the Korean Ophthalmological Society. 1999;40(10):2898–2903.

124. Li F, Kang J, Xu Y, Dong J, Qin X. Clinical Study of Highly Selective Dorsal Rhizotomy in Treating Spastic Cerebral Palsy. J Mod Med Health. 2000;(01):5–6.

125. Cao J, Guo X, He X. Exploration of cerebral palsy complication. Chinese J Rehabilitation Med. 2001;(01).

126. Yi B, Cao X, Jin Z. Functional outcome following selective posterior rhizotomy and analysis of the complications. Chinese J Rehabilitation Med. 2003;(04).

127. Jin C. Clinical Analysis of 31 Cases of Cerebral Palsy in Children. Proceedings of the 1st Conference of Child Rehabilitation of the Chinese Association of Rehabilitation Medicine. Published online 2004.

128. Chang F, Song J, Su Y. Superselective posterior rhizotomy for treatment of spastic cerebral palsy. Mod Med J Chin. 2006;(12):21–23.

129. Li F. Treatment of convulsionary brain paralysis with high alternative rhizotomy. Chin J Ortho Traumatol. 2006;(01):14–15.

130. Liao B, Lu W. The analysis of ametropia in 38 cerebral palsy (CP) patients. Beijing Med J. 2006;(12):712–714.

131. Wang X, Wang G, Liu J, Hu Y. Analysis of Strabismus in 220 Children with Cerebral Palsy. Chinese J Pract Pediatr. 2006;(07):545–546.

132. Liu Y, Liu Z. Analysis on visual function impediment in children with cerebral palsy. Chinese J Child Health Care. 2007;(01):71–72.

133. Liu Y, Gao L. Analysis of Strabismus in Children with Cerebral Palsy. J Med Forum. 2007;(09):92+94.

134. Wang S, Chen Y, Liu H. Effects of Carotid Sympathectomy on Intercurrent Damage of Refractory Cerebral Palsy: 204 Cases Report. Chinese J Rehabilitation Theor Pract. 2008;(09):858–859.

135. Li M. Analysis of Strabismus of Children with Cerebral Palsy. J Med Forum. 2009;(11):22-23+25.

136. Wu H, Sun G, Liu M, Ling X, Duan P, Cao J. Efficacy of Highly Selective Dorsal Rhizotomy in 346 Cases of Spastic Cerebral Palsy. Practical Clin Med. 2009;(11):67+74.

137. Fan Z. Preliminary Observation of Ocular Comorbidities in Children with Cerebral Palsy. Health Vocational Educ. 2010;(14):152–154.

138. Zhang X. Clinical Analysis on Infantile Cerebral Palsy in 55 Cases. J Henan Univ Sci Technol Med Sci. 2010;(03):202–203.

139. Li Y. Analysis on 208 cases of children with cerebral palsy. Master’s thesis, Jilin University. Published online 2011.

140. Marasini S, Paudel N, Adhikari P, JB S, Bowan M. Ocular Manifestations in Children With Cerebral Palsy. Optom Vis Devel. 2011;42:178–182.

141. Zhao J. Visual Dysfunction in Children with Cerebral Palsy. Master’s thesis, Qingdao University. Published online 2011.

142. Woo SJ, Ahn J, Park MS, et al. Ocular findings in cerebral palsy patients undergoing orthopedic surgery. Optom Vis Sci. 2011;88(12):1520–1523. doi:10.1097/OPX.0b013e3182346711

143. Wei H. Incidence of Strabismus in Cerebral Palsy. Proceedings of the 7th Beijing International Forum on Rehabilitation. Published online 2012.

144. Guo Y, Li Y, Yang Y, Peng Y, Zhao W. Analysis of eye examination of seventy-two children with spastic cerebral palsy. Int Eye Sci. 2013;(04):830–831.

145. Liu B. SPR combined carotid artery sympathetic nerve net stripping treatment efficacy analysis of mixed cerebral palsy. Master’s thesis, Xinjiang Medical University. Published online 2013.

146. Chen Y, Chen X. Analysis of Visual Function Abnormalities in 56 Children with Cerebral Palsy. Abstracts of the 14th International Congress of Ophthalmology and Optometry China;(COOC2014), The 3rd Congress of International Academy of Orthokeratology(CIAO2014). Published online 2014.

147. Luo Y, Tang J, Tan Y, Deng Z, Xiao Z, Tao L. Clinical research on visual impairment of children with cerebral palsy. Int Eye Sci. 2014;(04):782–784.

148. Lew H, Lee H, Lee J, Song J, Min K, Kim M. Possible linkage between visual and motor development in children with cerebral palsy. Pediatr Neurol. 2015;52(3):338–43.e1. doi:10.1016/j.pediatrneurol.2014.11.009

149. Wang P, Zhang H, Qin R, Tang J, Luo Y. Comprehensive visual impairment evaluation for cerebral palsy children. Int Eye Sci. 2015;(01):174–177.

150. Li X, Peng Q, Tian Y, Wu Q. Clinical analysis on visual impairment of children with cerebral palsy. Int Eye Sci. 2016;(02):392–394.

151. Lin B, Xu G, Liu J. A research on visual dystopia in 125 children with cerebral palsy. Chinese J Rehabilitation Med. 2016;(09):979–983.

152. Yan Y, Yu Z, Li L. Analysis of Risk Factors and Preventive Measures for Cerebral Palsy in Children with Premature Brain Injury. Syst Med. 2020;(18):147–149.

153. Govind A, Lamba PA. Visual disorders in cerebral palsy. Indian Journal of Ophthalmology. 1988;36(2):88.

154. Jethani J. Ocular defects in children with cerebral palsy. Indian J Ophthalmol. 2007;55(5):397.

155. Sasmal NK, Maiti P, Mandal R, et al. Ocular manifestations in children with cerebral palsy. J Indian Med Assoc. 2011;109(5):318, 323.

156. Vasanth J, Jacob N, Viswanathan S. Visual Function Status in Children with Cerebral Palsy. Optometry & Visual Performance. 2014;2(3).

157. Kaur G, Thomas S, Jindal M, Bhatti SM. Visual Function and Ocular Status in Children with Disabilities in Special Schools of Northern India. J Clin Diagn Res. 2016;10(10):NC01–NC04. doi:10.7860/JCDR/2016/23615.8742

158. Joshi MS, Telang OJ, Morepatil VG. OCULAR DISORDERS IN CHILDREN WITH DEVELOPMENTAL DELAY. jebmh. 2017;4(68):4065–4070. doi:10.18410/jebmh/2017/811

159. Khandaker. Associated impairments among children with cerebral palsy in rural Bangladesh: findings from the Bangladesh Cerebral Palsy Register (BCPR). Developmental Medicine & Child Neurology. 2017;59(S3):117–118. doi:10.1111/dmcn.63_13512

160. Mustafa J, Chaudhary A. Neuro-ophthalmological disorders in cerebral palsy and cortical visual impairment patients. Ophthalmology Pakistan. 2017;7(04):10–14.

161. Giliyar SK, Abid AR, Wagganavar PB, Kulkarni VM. Visual assessment in delayed development and cerebral palsy children in tertiary care center. Indian Journal of Clinical and Experimental Ophthalmology. 2018;4(1):67–69.

162. Raj R, Kotian VB. Ocular Evaluation of Patients with Cerebral Palsy. Acta Sci Ophthalmol. 2019;2:02–07.

163. Bhati P, Sharma S, Jain R, et al. Cerebral Palsy in North Indian Children: Clinico-etiological Profile and Comorbidities. J Pediatr Neurosci. 2019;14(1):30–35. doi:10.4103/jpn.JPN_46_18

164. Shaikh R. A Study on Ocular Manifestations Seen in Patients with Cerebral Palsy. JOAR. 2020;01(01). doi:10.46889/JOAR.2020.1105

165. Hassan S, Ali L, Alim Shaikh A, Shakila Sultana K, Karim R, Siddiqua A. Ocular Findings in Children with Cerebral Palsy. IOSR J Dent Med Sci (IOSR-JDMS). 2020;19(6):32–37.

166. Moly K, Choudhury M, Ahsan S, et al. Pattern of Squint (Strabismus) in Children with Cerebral Palsy – A Study Conducted in the Ophthalmology Out-Patient Department in a Tertiary Level Teaching Hospital in Bangladesh | Bioresearch Communications. BioRes Comm (BRC) [Internet]. 2019;5(1):655–658.

167. Mishra M, Adhikari K, Panigrahi N. A Prospective Cohort Study of Auditory and Visual Comorbidities in Children with Cerebral Palsy : Journal of Marine Medical Society. J Mar Med Soc. 2023;25:43–47.

168. Viswanath M, Jha R, Gambhirao AD, et al. Comorbidities in children with cerebral palsy: a single-centre cross-sectional hospital-based study from India. BMJ Open. 2023;13(7):e072365. doi:10.1136/bmjopen-2023-072365

169. Bala I, Tuli R, Dhiman I, Sharma R, Gautam P. A Study of Ocular Abnormalities in Children with Cerebral Palsy. DOS Times. 2023;29(2):57–61.

170. Bodunde OT, Popoopla DSA, Ojuawo A, Adeboye M a. N. OCULAR FINDINGS IN CHILDREN WITH CEREBRAL PALSY ATTENDING A TERTIARY HOSPITAL IN NORTH CENTRAL NIGERIA. Sierra Leone Journal of Biomedical Research. 2015;7(2):1–7. doi:10.4314/sljbr.v7i2

171. Duke R. Assessment and Management of Visual Perceptual Problems in Children with Cerebral Palsy in Cross River State, Nigeria. PhD (research paper style) thesis. London School of Hygiene & Tropical Medicine; 2021. doi:10.17037/PUBS.04664165

172. Duke RE, Nwachukuw J, Torty C, et al. Visual impairment and perceptual visual disorders in children with cerebral palsy in Nigeria. Br J Ophthalmol. 2022;106(3):427–434. doi:10.1136/bjophthalmol-2020-317768

173. Owunna C, Ekenze C, Okorie I, et al. Oculo-visual Assessment of Children and Adolescents with Special Needs in Selected Schools within IMO State, Nigeria. Ophthalmol Res: An Int J. 2022;16(3):8–19.

174. Musiwa M, Muma K, Mutoloki E. Ocular Evaluation in Cerebral Palsy Patients at Children’s and Eye Hospitals at the University Teaching Hospitals, Lusaka, Zambia. Journal of Advances in Medicine and Medical Research. 2023;35(4):1–7. doi:10.9734/jammr/2023/v35i44957

175. New World Bank country classifications by income level: 2022-2023. Published July 1, 2022. Accessed December 31, 2023. https://blogs.worldbank.org/opendata/new-world-bank-country-classifications-income-level-2022-2023

176. Higgins JPT, Thompson SG. Quantifying heterogeneity in a meta-analysis. Stat Med. 2002;21(11):1539–1558. doi:10.1002/sim.1186

177. DerSimonian R, Laird N. Meta-analysis in clinical trials. Controlled Clinical Trials. 1986;7(3):177–188. doi:10.1016/0197-2456(86)90046-2

178. Egger M, Smith GD, Phillips AN. Meta-analysis: principles and procedures. BMJ. 1997;315(7121):1533–1537. doi:10.1136/bmj.315.7121.1533

179. Striber N, Vulin K, Đaković I, et al. Visual impairment in children with cerebral palsy: Croatian population-based study for birth years 2003-2008. Croat Med J. 2019;60(5):414–420. doi:10.3325/cmj.2019.60.414

180. Banerjee TK, Hazra A, Biswas A, et al. Neurological disorders in children and adolescents. Indian J Pediatr. 2009;76(2):139–146. doi:10.1007/s12098-008-0226-z

181. El-Tallawy HN, Farghaly WM, Shehata GA, et al. Cerebral palsy in Al-Quseir City, Egypt: prevalence, subtypes, and risk factors. Neuropsychiatr Dis Treat. 2014;10:1267–1272. doi:10.2147/NDT.S59599

182. Okan N, Okan M, Eralp O, Aytekin AH. The prevalence of neurological disorders among children in Gemlik (Turkey). Dev Med Child Neurol. 1995;37(7):597–603. doi:10.1111/j.1469-8749.1995.tb12048.x

183. Foundation CPAR. Cerebral Palsy Facts | Cerebral Palsy Alliance Research Foundation. Accessed December 14, 2023. https://cparf.org/what-is-cerebral-palsy/facts-about-cerebral-palsy/

184. Jarvis S, Glinianaia SV, Arnaud C, et al. Case gender and severity in cerebral palsy varies with intrauterine growth. Arch Dis Child. 2005;90(5):474–479. doi:10.1136/adc.2004.052670

185. Taft LT. Cerebral palsy. Pediatr Rev. 1995;16(11):411–418; quiz 418.

186. Dowding VM, Barry C. Cerebral palsy: social class differences in prevalence in relation to birthweight and severity of disability. J Epidemiol Community Health. 1990;44(3):191–195. doi:10.1136/jech.44.3.191

187. Yeargin-Allsopp M, Van Naarden Braun K, Doernberg NS, Benedict RE, Kirby RS, Durkin MS. Prevalence of cerebral palsy in 8-year-old children in three areas of the United States in 2002: a multisite collaboration. Pediatrics. 2008;121(3):547–554. doi:10.1542/peds.2007-1270

188. Eunson P. Aetiology and epidemiology of cerebral palsy. Paediatrics and Child Health. 2016;26(9):367–372. doi:10.1016/j.paed.2016.04.011

189. Brodsky MC, Fray KJ, Glasier CM. Perinatal cortical and subcortical visual loss: mechanisms of injury and associated ophthalmologic signs. Ophthalmology. 2002;109(1):85–94. doi:10.1016/s0161-6420(01)00849-1

190. Ganesh S, Khurana R, Sharma S, Rath S. Predisposing Factors, Ophthalmic Manifestations, and Radiological Findings in Children With Cerebral Visual Impairment. J Pediatr Ophthalmol Strabismus. 2019;56(5):313–318. doi:10.3928/01913913-20190610-01

191. Sharbini S. Prevalence of strabismus and associated risk factors: the Sydney childhood eye studies. Dissertation, University of Sydney, Faculty of Health Sciences (Orthoptic). Published online 2015:290 pp.

192. Cioni G, Di Paco MC, Bertuccelli B, Paolicelli PB, Canapicchi R. MRI findings and sensorimotor development in infants with bilateral spastic cerebral palsy. Brain Dev. 1997;19(4):245–253. doi:10.1016/s0387-7604(97)00569-x

193. Wu YW, Xing G, Fuentes-Afflick E, Danielson B, Smith LH, Gilbert WM. Racial, ethnic, and socioeconomic disparities in the prevalence of cerebral palsy. Pediatrics. 2011;127(3):e674–681. doi:10.1542/peds.2010-1656

194. Lang TC, Fuentes-Afflick E, Gilbert WM, Newman TB, Xing G, Wu YW. Cerebral palsy among Asian ethnic subgroups. Pediatrics. 2012;129(4):e992–998. doi:10.1542/peds.2011-2283

195. Durkin MS, Maenner MJ, Benedict RE, et al. The role of socio-economic status and perinatal factors in racial disparities in the risk of cerebral palsy. Dev Med Child Neurol. 2015;57(9):835–843. doi:10.1111/dmcn.12746

196. Donnelly UM. Horizontal strabismus worldwide--what are the risk factors? Ophthalmic Epidemiol. 2012;19(3):117–119. doi:10.3109/09286586.2012.681002

197. Yang Y, Wang C, Gan Y, et al. Maternal smoking during pregnancy and the risk of strabismus in offspring: a meta-analysis. Acta Ophthalmol. 2019;97(4):353–363. doi:10.1111/aos.13953

198. Wiesinger H. Ocular Findings In Mentally Retarded Children. Journal of Pediatric Ophthalmology & Strabismus. 1964;1(3):37–41. doi:10.3928/0191-3913-19640701-08

199. Guzzetta A, Mercuri E, Cioni G. Visual disorders in children with brain lesions: 2. Visual impairment associated with cerebral palsy. Eur J Paediatr Neurol. 2001;5(3):115–119. doi:10.1053/ejpn.2001.0481

200. Sandfeld Nielsen L, Skov L, Jensen H. Visual dysfunctions and ocular disorders in children with developmental delay. II. Aspects of refractive errors, strabismus and contrast sensitivity. Acta Ophthalmol Scand. 2007;85(4):419–426. doi:10.1111/j.1600-0420.2007.00881.x

201. Plesca DA, Luhovschi C, Plesca VS, Cavache A. Prevalence of visual disturbances in children with spastic cerebral palsy. Intensive Care Medicine. 2011;37 SUPPL. 2:S383.

202. Mrugacz M, Bandzul K. Choroba zezowa u pacjentów z mózgowym porażeniem dziecięcym. Neurol Dziec. 2012;21(43):79–83.

203. Collins MLZ. Strabismus in cerebral palsy: when and why to operate. Am Orthopt J. 2014;64:17–20. doi:10.3368/aoj.64.1.17

204. Holm S. Le Strabisme Concomitant Chez Les Palénégrides Au Gabon, Afrique Equatoriale Française. Acta Ophthalmologica. 1939;17(4):367–387. doi:10.1111/j.1755-3768.1939.tb07376.x

205. Waardenburg PJ. Squint and heredity. Doc Ophthalmol. 1954;7-8:422–494. doi:10.1007/BF00238145

206. Laughton SC, Hagen MM, Yang W, von Bartheld CS. Gender differences in horizontal strabismus: Systematic review and meta-analysis shows no difference in prevalence, but gender bias towards females in the clinic. J Glob Health. 13:04085. doi:10.7189/jogh.13.04085

207. Agarwal AB, Feng CY, Altick AL, et al. Altered Protein Composition and Gene Expression in Strabismic Human Extraocular Muscles and Tendons. Invest Ophthalmol Vis Sci. 2016;57(13):5576–5585. doi:10.1167/iovs.16-20294

208. Jacobson LK, Dutton GN. Periventricular leukomalacia: an important cause of visual and ocular motility dysfunction in children. Surv Ophthalmol. 2000;45(1):1–13. doi:10.1016/s0039-6257(00)00134-x

209. McDonnell K, O’Connor AR. The significance of periventricular leukomalacia on ophthalmic outcome. British and Irish Orthoptic Journal. 2015;12:2–8.

210. Tychsen L, Lisberger SG. Maldevelopment of visual motion processing in humans who had strabismus with onset in infancy. J Neurosci. 1986;6(9):2495–2508. doi:10.1523/JNEUROSCI.06-09-02495.1986

211. Tychsen L. The cause of infantile strabismus lies upstairs in the cerebral cortex, not downstairs in the brainstem. Arch Ophthalmol. 2012;130(8):1060–1061. doi:10.1001/archophthalmol.2012.1481

212. Jeon H, Jung J, Kim H, Yeom JA, Choi H. Strabismus in children with white matter damage of immaturity: MRI correlation. Br J Ophthalmol. 2017;101(4):467–471. doi:10.1136/bjophthalmol-2016-308769

213. Araneda R, Ebner-Karestinos D, Dricot L, et al. Impact of early brain lesions on the optic radiations in children with cerebral palsy. Front Neurosci. 2022;16:924938. doi:10.3389/fnins.2022.924938

214. Khanna S, Sharma A, Ghasia F, Tychsen L. Prevalence of the Infantile Strabismus Complex in Premature Children With and Without Periventricular Leukomalacia. Am J Ophthalmol. 2022;240:342–351. doi:10.1016/j.ajo.2022.03.028

215. Ganesh S, Rath S. Cerebral Visual Impairment in Children. Delhi Journal Of Ophthalmology. 2018;29. doi:10.7869/djo.389

216. Hiwale AJ, Chandra Das K. Geospatial differences among natural regions in the utilization of maternal health care services in India. Clinical Epidemiology and Global Health. 2022;14:100979. doi:10.1016/j.cegh.2022.100979

217. Reddihough DS, Collins KJ. The epidemiology and causes of cerebral palsy. Aust J Physiother. 2003;49(1):7–12. doi:10.1016/s0004-9514(14)60183-5

